# Single-cell profiling reveals monocyte mitochondrial dysfunction in patients with cirrhosis progressing to acute-on-chronic liver failure

**DOI:** 10.1101/2025.02.04.25321370

**Authors:** Theresa Hildegard Wirtz, Sara Palomino-Echeverria, Maike Rebecca Pollmanns, Estefanía Huergo, Felix Schreibing, Johanna Reißing, Cristina Sánchez-Garrido, Cristina López-Vicario, Ana Maria Aransay, Maurizio Baldassare, Giacomo Zaccherini, Enrico Pompili, Martin Schulz, Frank Erhard Uschner, Sabine Klein, Wenyi Gu, Robert Schierwagen, Shantha Valainathan, Annabelle Verbeeck, Daniela Campion, Ilaria Giovo, Kai Markus Schneider, Alexander Koch, Rafael Kramann, Tony Bruns, Narsis Kiani, Linsey Stiles, Paolo Caraceni, Carlo Alessandria, Richard Moreau, Jonel Trebicka, Joan Clària, Núria Planell, Pierre-Emmanuel Rautou, Christian Trautwein, David Gomez-Cabrero

## Abstract

**Background & Aims:** Patients with acute decompensation (AD) of cirrhosis are at a high risk of developing acute-on-chronic liver failure (ACLF), a syndrome characterized by multi-organ failure and high short-term mortality. Prospective studies addressing the cellular mechanisms that drive the transition from AD to ACLF are lacking. In this study, we aimed to determine whether peripheral immune cell subsets at hospital admission predict the progression from AD to ACLF and to delineate underlying molecular mechanisms.

**Approach & Results:** We prospectively enrolled 63 patients with AD and 15 healthy donors across five European centers, with a 90-day follow-up. Single-cell RNA sequencing was performed on peripheral blood mononuclear cells (PBMCs) from 16 patients with distinct trajectories and 4 controls. Progression to ACLF was associated with expansion of classical monocytes, particularly subcluster “C2”, which specifically displayed impaired energy metabolism with reduced oxidative phosphorylation. Genes encoding respiratory chain Complex IV were markedly downregulated. A C2-derived gene signature was enriched in two large international whole-blood cohorts (PREDICT, n=689; ACLARA, n=521), particularly in patients with bacterial infections, those developing ACLF, and non-survivors. Functional validation by respirometry in an independent AD cohort confirmed declining monocyte Complex IV–dependent oxygen consumption in patients with pre-ACLF.

**Conclusions:** We identified and validated a distinct monocyte subpopulation with defective energy metabolism in patients with AD with poor outcomes, suggesting a mechanistic link to ACLF development.

## INTRODUCTION

Acute decompensation (AD) is the most frequent cause of hospitalization among patients with cirrhosis (1). It is characterized by the development of clinically relevant complications, including ascites, hepatic encephalopathy, or gastrointestinal bleeding (2–4). Patients with AD are at high risk of progressing to acute-on-chronic liver failure (ACLF), a syndrome defined by acute deterioration of liver function accompanied by failure of one or more extrahepatic organs (5,6). Depending on the number and severity of organ failures, ACLF is associated with short-term mortality rates ranging from 15% to 80% (7).

The transition from decompensated cirrhosis to ACLF is a dynamic and multifactorial process driven by systemic inflammation, immune dysfunction, and organ crosstalk (8,9). Systemic inflammation progressively increases with worsening portal hypertension and contributes to organ failure during ACLF development (10,11). Elevated circulating pro-inflammatory cytokines, activation of innate immune pathways such as Toll-like receptor signaling, and translocation of microbial products due to intestinal barrier dysfunction have been identified as key drivers of this process (12,13). In parallel, immune cells undergo phenotypic and functional alterations that impair host defense and increase susceptibility to bacterial infections, a major precipitating factor of ACLF (14,15).

Despite these advances, the relative contribution of individual immune cell subsets to disease progression remains incompletely defined. In particular, it is unclear which functional immune cell states precede and predict the development of ACLF in patients with AD. Single-cell RNA sequencing (scRNA-seq) enables high-resolution characterization of immune cell heterogeneity and has recently been applied to patients with established ACLF, revealing distinct immunometabolic alterations, particularly in monocytes and T cells (16–19). However, these studies focused on patients who had already transitioned to ACLF. Prospective high-resolution analyses of peripheral immune cell alterations in AD patients who subsequently develop ACLF are lacking.

Here, we conducted a prospective multicenter study that included patients admitted with AD across five European centers. Using single-cell transcriptomic profiling of peripheral blood mononuclear cells (PBMCs), we aimed to characterize immune cell states associated with disease progression. Our data provide insight into dynamic changes in peripheral immune cell subpopulations and identify molecular mechanisms linked to the progression from AD to ACLF.

## MATERIAL AND METHODS

### Study design

We conducted a prospective multicenter study at five European centers (Paris, Frankfurt, Aachen, Bologna, and Turin) between October 2020 and July 2021. Patients admitted to the hospital due to AD (ascites, overt hepatic encephalopathy, new onset of non-obstructive jaundice, gastrointestinal hemorrhage, and/or bacterial infections) met the inclusion criterion and were screened for exclusion criteria (**Table S1**). A total of 63 patients with AD and 15 healthy controls were recruited for this study. Clinical data were collected, and blood draw was performed within 72 h after hospital admission in all included patients. Patients were followed up for 90 days, and readmission due to AD, development of ACLF, and outcomes were documented (for a detailed study design and visit schedule, **Fig. S1** and **Table S2**).

### Ethics

Ethics approval was obtained at each center before patient enrollment and name of the ethics committees and number/ID of the approvals were as follows: Paris (“Comite de protection de personnes”): 2016-A01599-42, Frankfurt (“Ethikkommission des Fachbereichs Medizin der Goethe-Universität Frankfurt am Main”): 20-653; Aachen (“Ethik-Kommission an der Medizinischen Fakultät der RWTH Aachen”): EK024/19; EK069/16, Bologna (“Comitato Etico Area Vasta Emilia Centro”): 8/2017/U/Tess, Turin (“Comitato Etico Interaziendale”): CS2/181.

### Group allocation and patient selection strategy for single-cell analysis

90 days after study inclusion, patients were classified according to their clinical course as no readmission due to AD (“stable decompensated cirrhosis”, SDC, n=46), readmission due to AD (“unstable decompensated cirrhosis”, UDC, n=8), or ACLF development within 28 days (“pre-ACLF”, n=9). The definition of SDC and UDC was pre-specified before study initiation in accordance with Trebicka J. *et al (7)*. The diagnostic criteria of ACLF were according to the EF-CLIF definition and included the presence of organ failure (5). Of the 63 recruited patients, n=9 developed ACLF within 90 days. Of those, 5 patients were excluded from CITE-seq analysis due to one of the following reasons: “late ACLF development” (>28 days post-hospital admission), “incomplete 90d follow-up” or “cells not viable”. The 4 remaining samples were included in CITE-seq analysis (**Table S3**). Subsequently, 6 samples from the SDC and UDC group each as well as 4 healthy controls were selected for CITE-seq ensuring a balance of demographic and clinical characteristics between the four study groups (**Table S4**). Table S5 depicts a detailed overview on readmission(s) and outcome of each included patient (**Table S5**).

### Further methods

Detailed information on sample processing, single-cell and bulk RNA sequencing, flow cytometry, functional assays, validation cohorts, statistical analyses, and data availability is provided in the **Supplementary Methods**.

## RESULTS

### Study design and baseline characteristics of the single-cell study cohort

In total, 63 patients with AD and 15 healthy controls were prospectively recruited across five European centers. After 90 days of follow-up, patients were classified as stable decompensated cirrhosis (SDC), unstable decompensated cirrhosis (UDC), or pre-ACLF (development of ACLF within 28 days). Sixteen patients (6 SDC, 6 UDC, 4 pre-ACLF) and four healthy controls were selected for single-cell analysis (s. Material & Methods, **Fig. 1A**). PBMCs underwent CITE-seq, and matched whole-blood RNA-seq was performed in the same individuals (**Fig. 2A**). Patients in the pre-ACLF exhibited significantly higher MELD and CLIF-C AD scores compared with SDC and UDC patients (**Table 1**). Bacterial infections were more frequent in pre-ACLF, including spontaneous bacterial peritonitis. Treatment at the time of blood draw was not significantly different between the groups. Detailed baseline characteristics and follow-up data are provided in **Table 1** and **Tables S3–S5**.

**Table 1.**
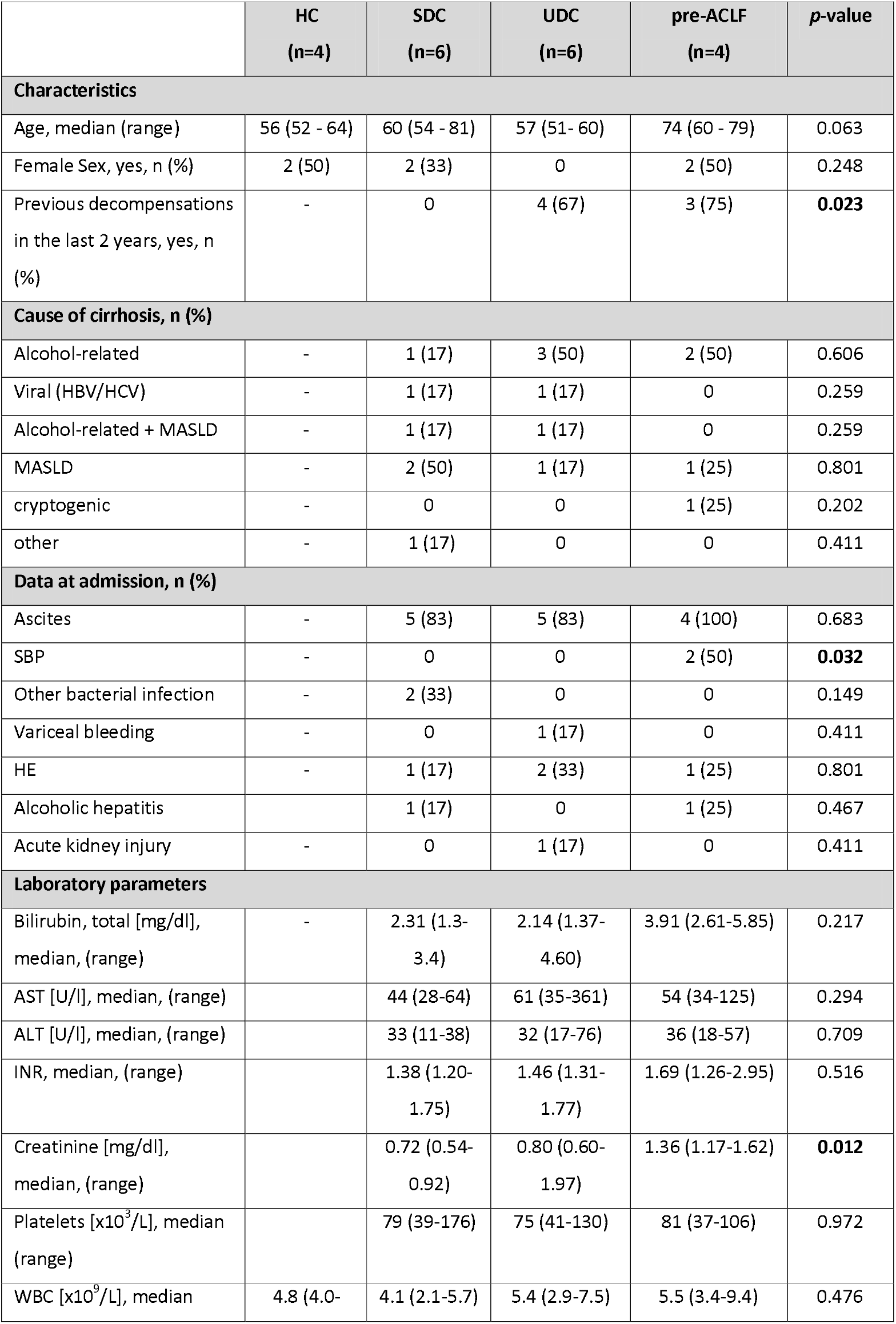
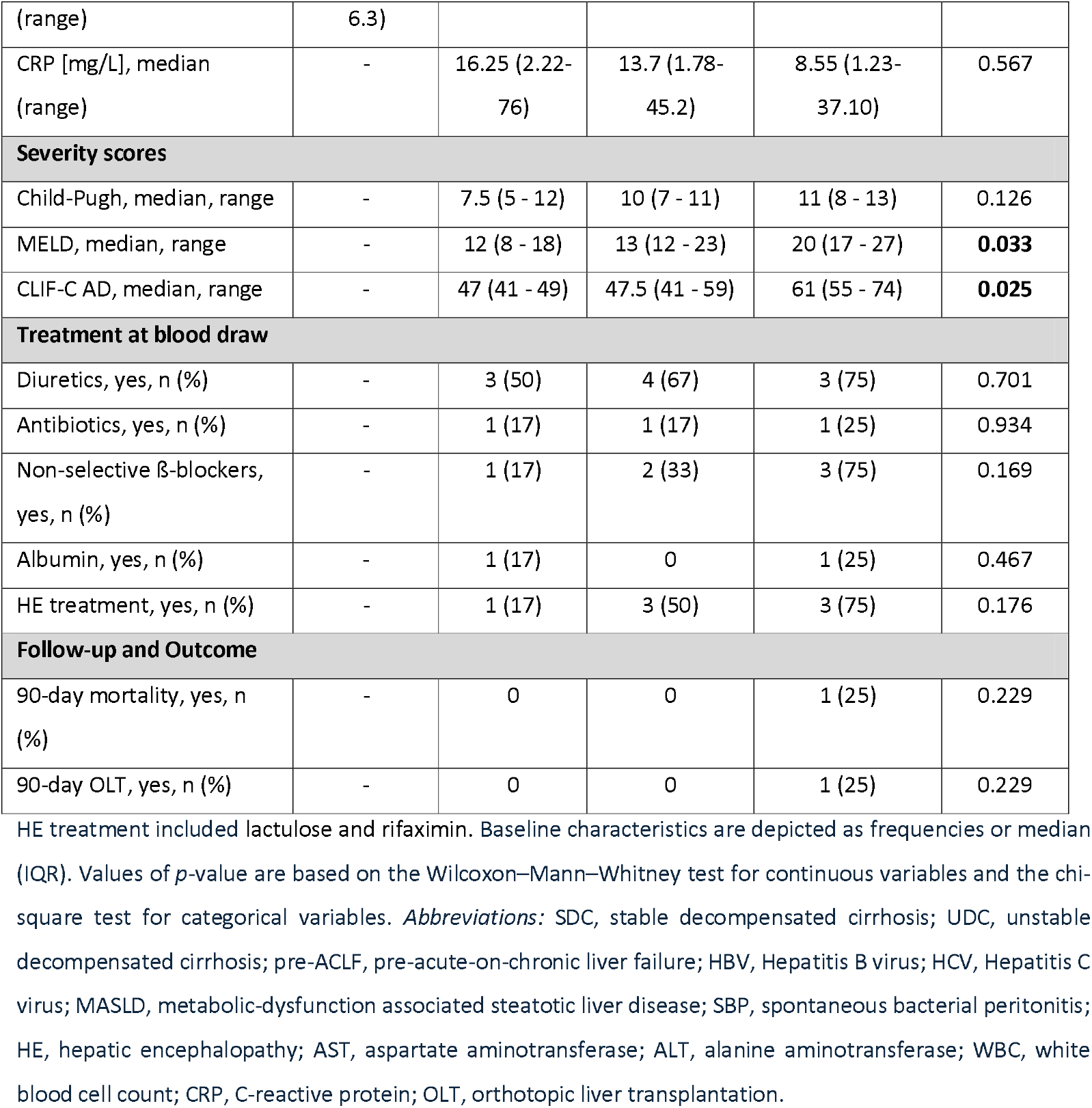
Baseline characteristics of single-cell study cohort.

**Fig. 1.**
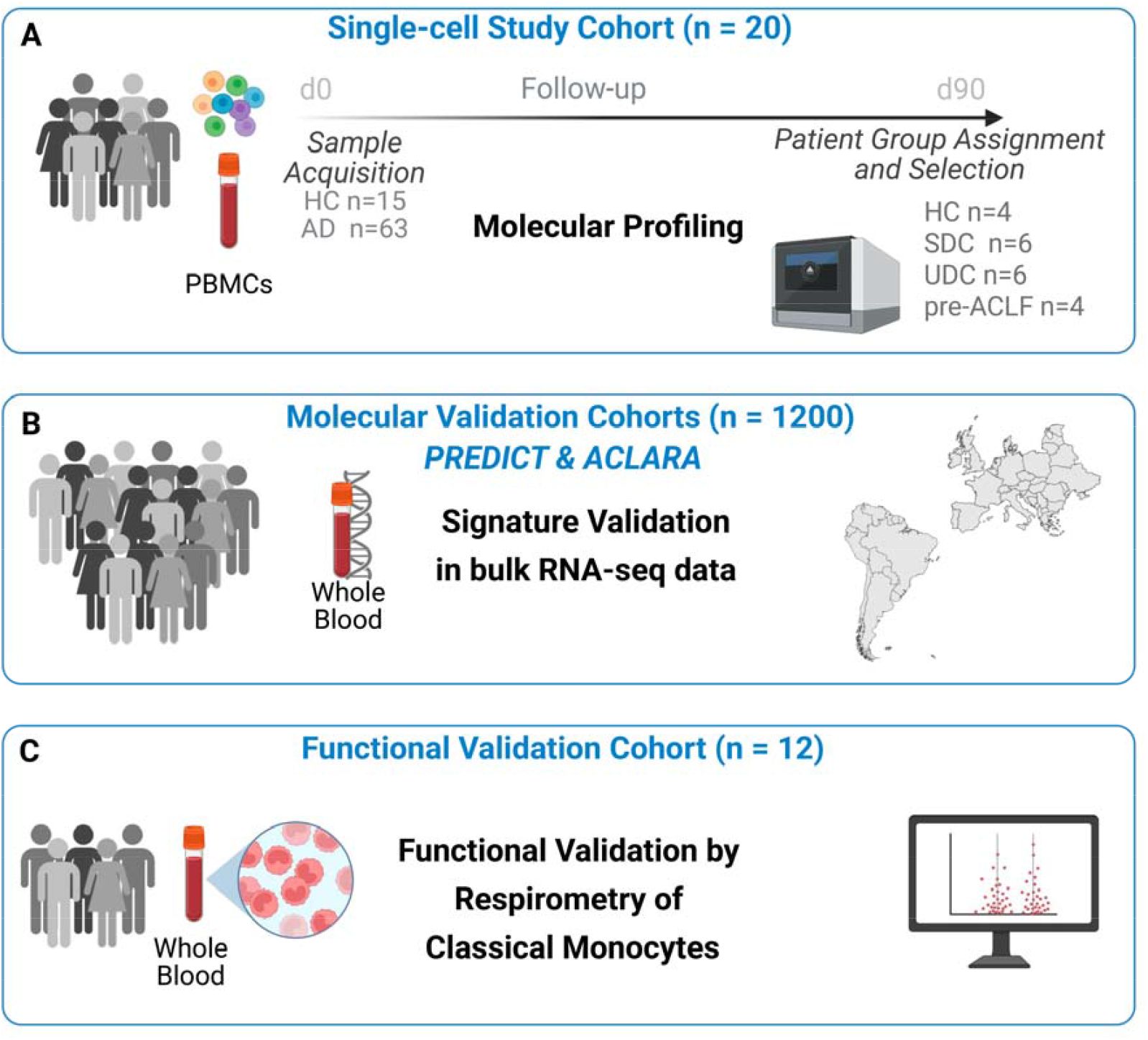
Overview of study cohorts and analytical workflow. Single-cell profiling **(A)**, whole-blood transcriptomic validation **(B)**, and functional validation cohorts **(C)** were analyzed in this study. *Abbreviations:* AD, acute decompensation; HC, healthy controls; PBMCs, peripheral blood mononuclear cells; pre-ACLF, pre–acute-on-chronic liver failure; SDC, stable decompensated cirrhosis; UDC, unstable decompensated cirrhosis.

**Fig. 2.**
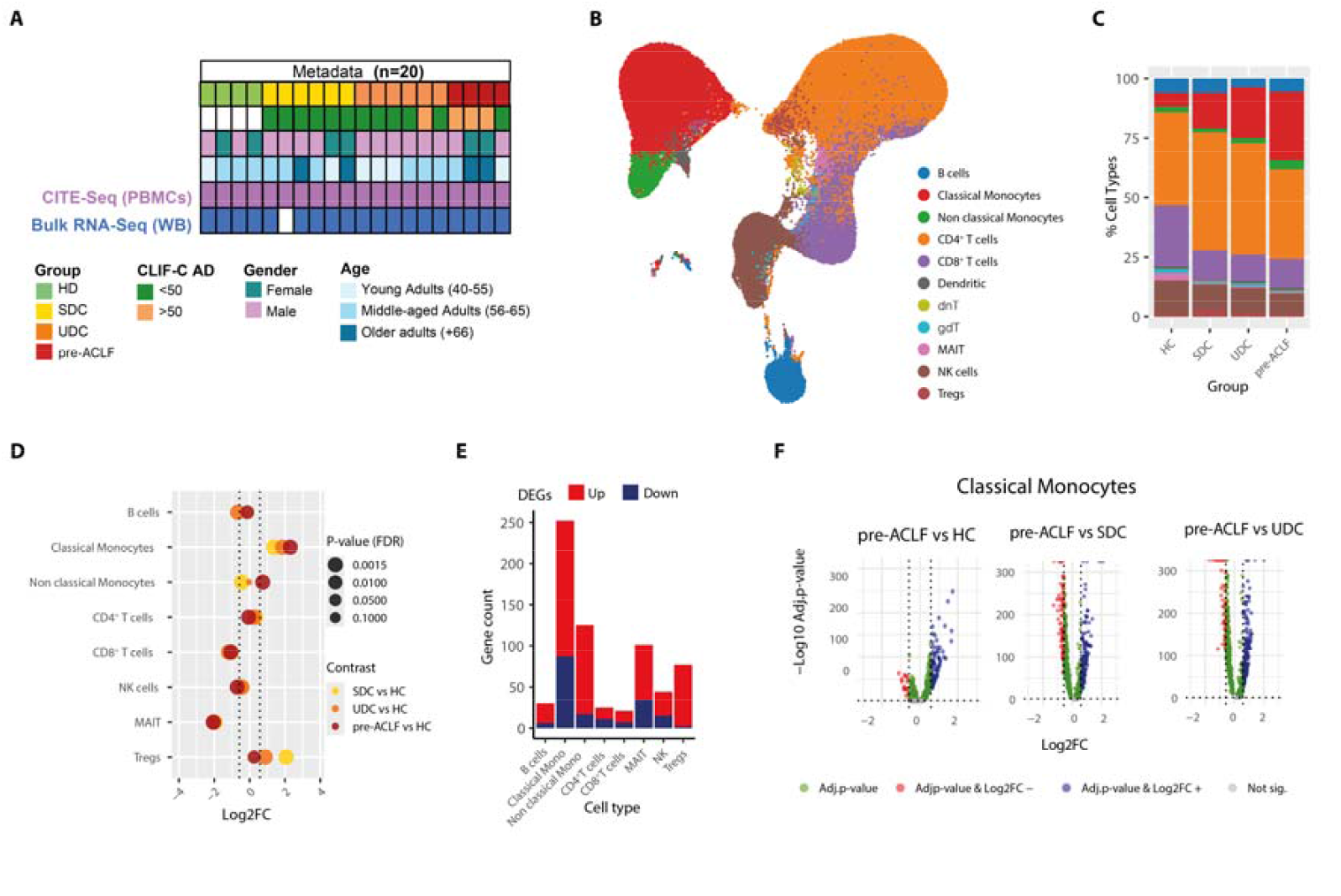
An increase in circulating classical monocytes is associated with ACLF development. **(A)** Overview of the multi-omics study design and associated clinical metadata, including study group, CLIF-C AD score, sex, and age. **(B)** UMAP projection of 143,952 PBMCs colored by the 11 identified immune cell populations. **(C)** Relative proportions of immune cell subsets across study groups. **(D)** Cleveland plot showing log_2_ fold changes of the eight major immune cell populations relative to healthy controls. Comparisons include SDC vs. HC (yellow), UDC vs. HC (orange), and pre-ACLF vs. HC (dark red). Cell types were considered differentially abundant when log_2_ fold change > 0.58 and adjusted p < 0.05. **(E)** Total number of differentially expressed genes (DEGs) per cell type. DEGs were defined by adjusted p < 0.05 (Benjamini–Hochberg) and absolute fold change > 1.5. **(F)** Volcano plots showing differential gene expression in classical monocytes comparing pre-ACLF, UDC, and SDC patients with healthy controls. The x-axis indicates log2 fold change and the y-axis −log_10_ adjusted p-value. Red and blue dots indicate significantly up- or downregulated genes; grey dots indicate non-significant genes. *Abbreviations:* AD, acute decompensation; HC, healthy controls; MAIT, mucosal-associated invariant T cells; NK, natural killer cells; PBMCs, peripheral blood mononuclear cells; pre-ACLF, pre-acute-on-chronic liver failure; SDC, stable decompensated cirrhosis; Tregs, regulatory T cells; UDC, unstable decompensated cirrhosis.

### An increase in circulating classical monocytes is associated with ACLF development

For the single-cell transcriptional analysis, 130,846 cells, with a median of 1,632 genes per cell, were recovered from the 20 individuals included in this study **(Fig. 1A)**. Unsupervised clustering identified 11 immune cell populations, including classical and non-classical monocytes, CD4^+^ and CD8^+^ T cells, B cells, NK cells, MAIT cells, dendritic cells, regulatory T cells, dnT cells, and γδ T cells (**Fig. 2B, Fig. S2**). Cell-type annotation was confirmed by surface marker expression (**Fig. S3A, B**).

Comparison of cell-type proportions across healthy controls, SDC, UDC, and pre-ACLF patients revealed pronounced changes among the most abundant immune populations. Classical monocytes were significantly increased in AD patients compared to healthy controls, with the highest fold change observed in pre-ACLF, followed by UDC and SDC (**Fig. 2C, D**). Non-classical monocytes were also increased in pre-ACLF patients, albeit to a lesser extent, whereas CD8^+^ T cells were reduced in AD patients relative to healthy controls. Similar trends were observed using surface protein marker data (**Fig. S3C, D**).

Differential expression analysis across cell types identified marked transcriptional alterations in AD patients. Classical monocytes exhibited the highest number of differentially expressed genes (DEGs) across all contrasts when compared with healthy controls (**Fig. 2E**). Direct comparisons of pre-ACLF with SDC and UDC further confirmed that classical monocytes displayed the most pronounced transcriptional changes (**Fig. 2F; Fig. S4**).

Together, these analyses demonstrate substantial remodeling of the peripheral immune cell landscape in AD, characterized by a selective expansion and transcriptional activation of classical monocytes, most prominently in patients progressing to ACLF.

### Identification of a classical monocyte subpopulation associated with AD progression

Based on the observed alterations in abundance and gene expression of classical monocytes in AD patients, we performed subclustering of classical monocytes to identify disease-associated subsets. Clustering analysis identified three subclusters (C0–C2), which aligned along a pseudotime trajectory inferred using Monocle3, with C1 representing an intermediate state between C0 and C2 (**Fig. 3A, B**). C0 constituted the largest fraction of classical monocytes, followed by C1 and C2.

**Fig. 3.**
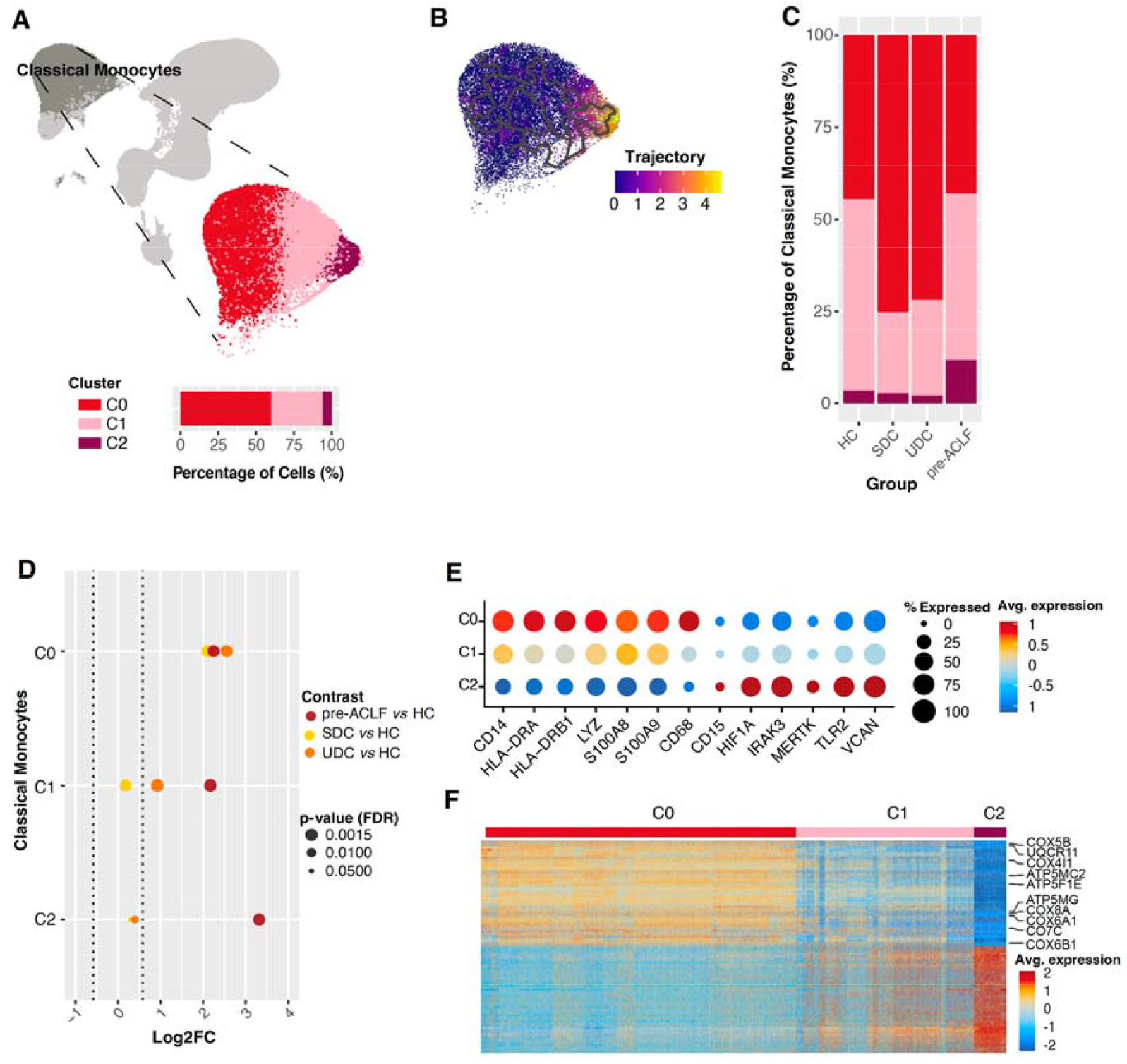
C2 monocytes exhibit a distinct immunoregulatory and metabolically repressed phenotype. **(A)** UMAP of PBMCs highlighting classical monocytes and their subdivision into three clusters (C0–C2). **(B)** Pseudotime trajectory of classical monocytes inferred using Monocle3. **(C)** Relative proportions of C0, C1, and C2 monocytes across study groups. **(D)** Log_2_ fold changes in subcluster abundance relative to healthy controls for SDC, UDC, and pre-ACLF patients. **(E)** Dot plot showing expression of selected marker genes across monocytic subclusters. **(F)** Heatmap of scaled expression values for the top differentially expressed genes across C0–C2 (see Fig. S7 for full annotation). *Abbreviations:* Avg, average; HC, healthy controls; Log_2_FC, log_2_ fold change; pre-ACLF, pre-acute-on-chronic liver failure; SDC, stable decompensated cirrhosis; UDC, unstable decompensated cirrhosis.

Analysis of subcluster distribution across study groups revealed that all three subclusters were expanded in AD patients compared to healthy controls (**Fig. 3C; Fig. S5A**). While C0 was increased across all AD groups, C1 was enriched in UDC and pre-ACLF patients, and C2 was selectively increased in pre-ACLF patients (**Fig. 3D**). Within the pre-ACLF group, the proportions of C1 and C2 increased with shorter latency to ACLF onset (**Fig. S5B**).

### C2 monocytes exhibit an immunoregulatory and metabolically repressed phenotype

Transcriptional profiling of monocytic marker genes revealed that C2 monocytes exhibited reduced expression of *CD14*, HLA-DR–encoding genes (*HLA-DRA, HLA-DRB1*), genes encoding for S100 family members, and the antibacterial protein lysozyme (*LYZ*) compared with C0 (**Fig. 3E**). In contrast, genes involved in immune regulation and inflammatory signaling, including *MERTK, VCAN, IRAK3, HIF1A*, and *TLR2*, were upregulated in C2, whereas C1 showed an intermediate expression profile.

Comparison of the C2 transcriptional program with a published endotoxin-tolerant monocyte signature (MS1) revealed a limited overlap and no significant enrichment of the MS1 gene set in C2 monocytes (**Fig. S6A–C**).

Differential expression analysis of C2 and C0/C1 confirmed the intermediate state of C1 (**Fig. 3F**; **Fig. S7**). Notably, genes involved in cellular energy metabolism were prominently downregulated in C2, including multiple components of ATP synthase (Complex V) and cytochrome c oxidase (Complex IV). In total, three genes encoding ATP synthase/Complex V membrane subunits and six encoding cytochrome c oxidase/Complex IV subunits were among the 50 most downregulated genes in the C2 subset compared to C0/C1 (**Fig. 3F**). Several S100 family genes were also among the most downregulated transcripts in C2 (**Fig. S7**).

Consistent with reduced HLA-DR transcript levels in the C2 subset, flow cytometric analysis of the entire population of classical monocytes enriched from PBMCs from the same AD and healthy control individuals showed a trend toward lower HLA-DR surface expression in pre-ACLF patients than in other groups (**Fig. S8**).

Together, these results identify a distinct classical monocyte subpopulation, whose expansion is associated with progression to ACLF and is characterized by immunoregulatory features and suppression of genes involved in cellular energy metabolism.

### C2 monocytes are defined by profound suppression of mitochondrial respiration

To better understand the biological functions of the identified monocyte subsets, we conducted gene set enrichment analysis (GSEA) to compare C2 and C0, with C1 as an intermediate state. Oxidative phosphorylation (OXPHOS) was the most negatively enriched pathway in C2 monocytes (**Fig. 4A; Table S6**). Consistently, OXPHOS module scores showed a graded decline across subclusters (C0 > C1 > C2) (**Fig. 4B**).

**Fig. 4.**
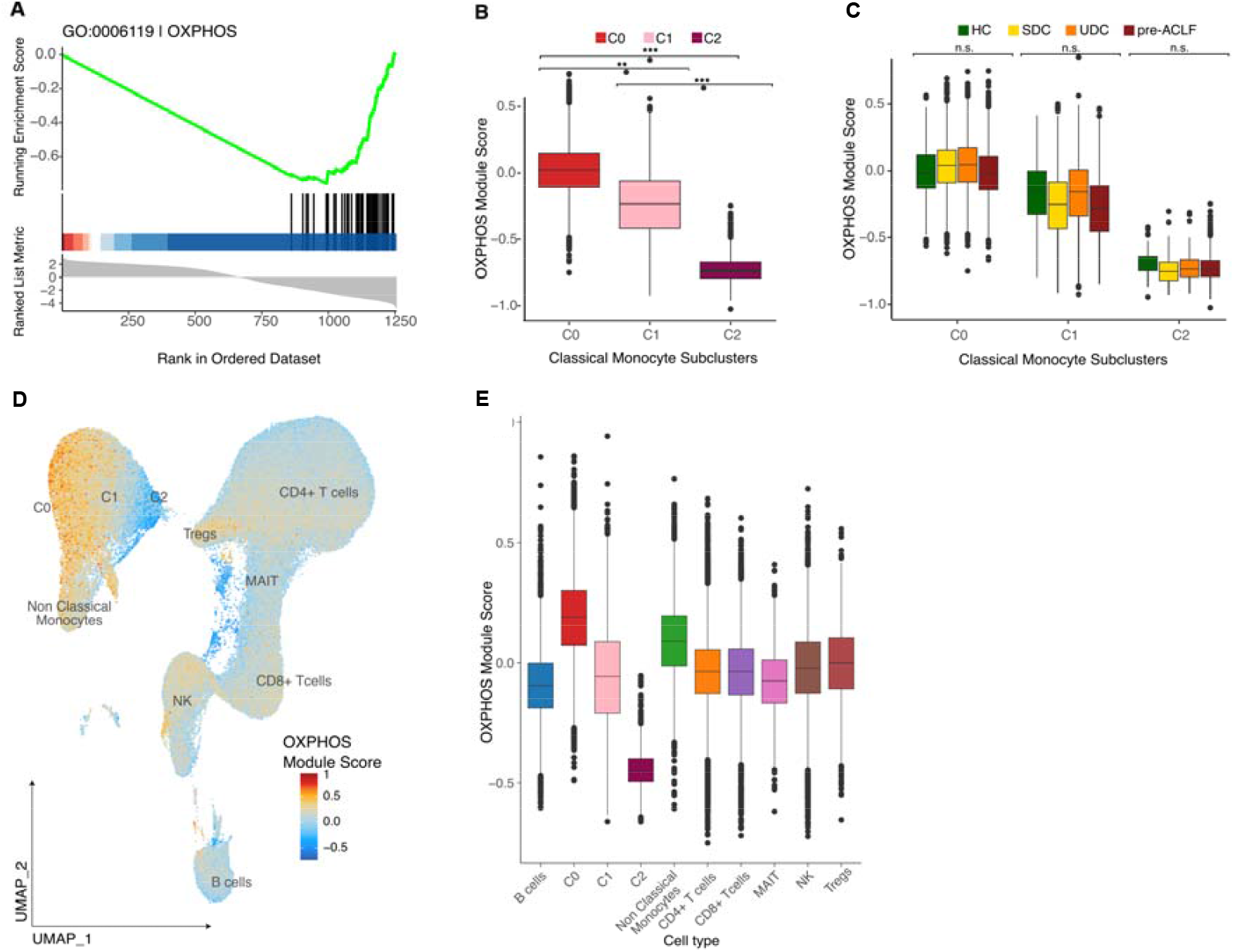
C2 monocytes are characterized by suppression of oxidative phosphorylation. **(A)** Gene set enrichment analysis (GSEA) identifying oxidative phosphorylation (OXPHOS) as a negatively enriched pathway in C2 monocytes. **(B)** OXPHOS module scores across classical monocyte subclusters. **(C)** OXPHOS module scores across monocyte subclusters stratified by clinical group. **(D)** UMAP projection of all cells colored by OXPHOS module score. **(E)** OXPHOS module scores across major immune cell types. Pairwise Wilcoxon rank-sum tests were used to assess differences in median scores; all pairwise comparisons were significant except CD8 vs CD4 T cells (ns). Statistical significance was assessed using Kruskal–Wallis or Wilcoxon rank-sum tests, as indicated. *** adjusted *p* < 0.001; ** adjusted *p* < 0.01; n.s., not significant. *Abbreviations:* HC, healthy controls; OXPHOS, oxidative phosphorylation; pre-ACLF, pre-acute-on-chronic liver failure; SDC, stable decompensated cirrhosis; UDC, unstable decompensated cirrhosis.

Stratification by clinical group further confirmed that C2 monocytes exhibited the lowest OXPHOS activity across disease stages (**Fig. 4C**). Importantly, the reduced OXPHOS activity was specific to the C2 subset among investigated immune cell subsets (**Fig. 4D, E; Fig. S9**). These findings indicate a robust and context-independent suppression of mitochondrial respiration in C2 monocytes.

Given the relevance of bacterial infection in ACLF progression, we additionally assessed immune-related pathways in C2 monocytes. Although no immune pathways reached statistical significance after multiple testing corrections, several showed a consistent trend toward reduced activity, including pathways related to inflammatory responses, antibacterial defense, and cytokine production (**Table S6**).

Together, these results demonstrate that C2 monocytes are characterized by profound impairment of cellular energy metabolism, accompanied by a tendency toward immune functional suppression.

### Identification and translation of a C2 monocytic gene signature to whole blood profiles

Based on the identification of an expanded C2 monocyte population in pre-ACLF patients and the availability of matched whole-blood transcriptomic data, we investigated whether a C2-associated gene signature could be detected at the systemic level (**Fig. 5A**). Using scRNA-seq data, we defined a C2 signature by selecting genes that were (i) upregulated in classical monocytes, (ii) enriched in C2 compared with other classical monocyte subsets, and (iii) specifically increased in C2 from pre-ACLF patients. This approach identified 96 genes that characterized the C2 subset in pre-ACLF patients.

**Fig. 5.**
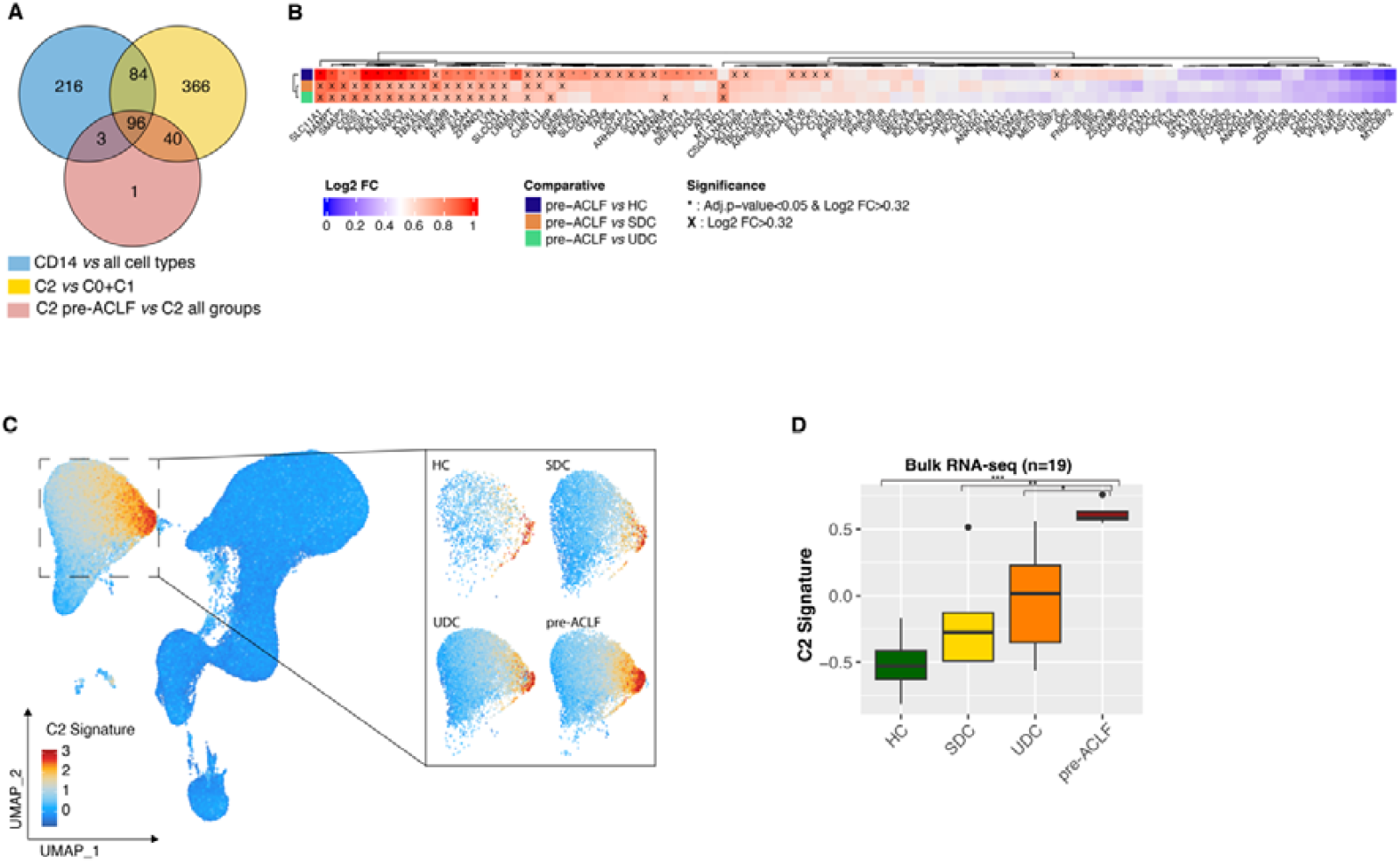
Identification and translation of a C2 gene signature to whole blood RNA-seq data of the single-cell study cohort. **(A)** Venn diagram showing the overlap of differentially expressed genes (DEGs) identified at single-cell resolution from three comparisons: classical monocytes versus all other cell types, C2 versus C0/C1, and C2 from pre-ACLF patients versus C2 from HC, SDC, and UDC. **(B)** Heatmap of scaled log_2_ fold changes of C2 signature genes in bulk RNA-seq data comparing pre-ACLF with HC, SDC, and UDC. Significant DEGs with log2 FC > 0.32 and adjusted p-value < 0.05 are marked with an asterisk; genes with a tendency (log2 FC > 0.32) are marked with a cross. **(C)** UMAP visualization of C2 signature activity at the single-cell level, with inset panels showing group-specific distributions. **(D)** C2 signature module scores in paired whole-blood RNA-seq samples stratified by clinical group. *Abbreviations:* HC, healthy controls; log_2_FC, log_2_ fold change; pre-ACLF, pre-acute-on-chronic liver failure; SDC, stable decompensated cirrhosis; UDC, unstable decompensated cirrhosis.

Analysis of these genes in whole-blood RNA-seq data showed that 94 genes were detectable at the bulk level. Applying a fold-change threshold in bulk RNA-seq identified 18 genes defining a C2 signature in whole blood: *SLC11A1, NAMPT, SMAP2, CD55, ACSL1, NEAT1, DLEU2, IRAK3, KYNU, TBXAS1, FKBP5, NUMB, PHF21A, AOAH, ZFAND3, LYN, SLCO3A1*, and *PTEN* (**Fig. 5B**).

To quantify the C2 signature activity, we computed a module score in both single-cell PBMC and whole-blood RNA-seq data (**Fig. 5C, D**). C2 signature activity progressively increased from healthy controls to SDC and UDC and was highest in pre-ACLF patients at both single-cell and bulk transcriptomic levels.

Together, these results demonstrate that the C2 monocytic transcriptional program identified at single-cell resolution can be robustly detected in whole blood and is enriched in patients progressing to ACLF.

### Independent cohort validation establishes clinical relevance of the C2 signature in patients with AD

To validate the clinical significance of the defined C2 gene signature, we evaluated its relevance in two large and independent prospective multicenter cohorts of patients with AD from Europe (PREDICT; n = 689) and Latin America (ACLARA; n = 521) (**Fig. 1B, Fig. 6A**). In both cohorts, whole-blood samples were collected at hospital admission and subjected to bulk RNA sequencing. In the PREDICT cohort, patients were followed for 90 days and classified as SDC, UDC, or pre-ACLF based on their clinical trajectory (**Fig. S10, Table S7**). In the ACLARA cohort, patients were followed up for 28 days and classified as AD or pre-ACLF (**Fig. S11, Table S8**). Detailed descriptions of patient selection are provided in the Materials and Methods section and **Fig. S12** and **S13**.

**Fig. 6.**
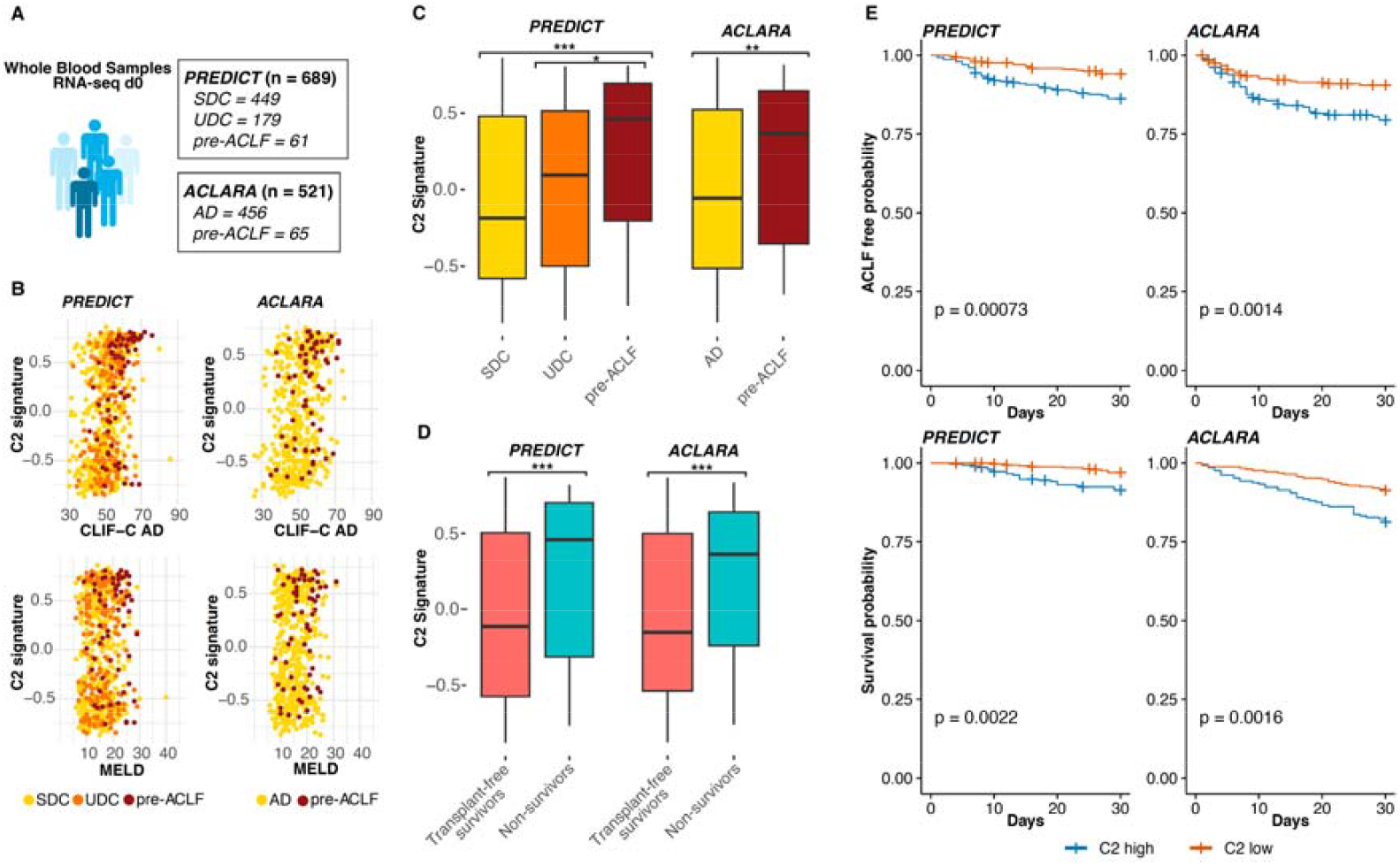
Independent cohort validation demonstrates clinical relevance of the C2 signature in patients with AD. **(A)**Assessment of the C2 signature in whole-blood RNA-seq data collected at hospital admission from two independent prospective cohorts: PREDICT (n = 689) and ACLARA (n = 521). **(B)** Correlation of C2 signature activity with CLIF-C AD score and MELD in the PREDICT and ACLARA cohorts. **(C)** C2 signature activity across clinical groups in PREDICT and ACLARA. **(D)** Comparison of C2 signature activity between survivors and non-survivors in both cohorts. **(E)** Kaplan–Meier analyses showing ACLF-free survival (top) and overall survival (bottom) stratified by C2 signature activity (high vs. low) in the PREDICT and ACLARA cohorts. Statistical significance was assessed using Wilcoxon rank-sum or log-rank tests, as indicated. *** adjusted *p* < 0.001; ** adjusted *p* < 0.01. *Abbreviations:* CLIF-C AD, Chronic Liver Failure Consortium Acute Decompensation score; MELD, Model for End-Stage Liver Disease; pre-ACLF, pre-acute-on-chronic liver failure; SDC, stable decompensated cirrhosis; UDC, unstable decompensated cirrhosis.

After computing the C2 signature activity score in both cohorts, we observed a significant positive correlation with the established indicators of liver disease severity, including total bilirubin, MELD, and CLIF-C AD score at hospital admission (**Fig. 6B**; s. **Tables S9 and S10** for correlation coefficients). Consistently, C2 signature activity was significantly higher in pre-ACLF patients and in non-survivors in both cohorts (**Fig. 6C, D**), supporting an association of the C2 monocyte program with disease severity, AD progression, and fatal outcome.

To further explore pathophysiological correlates of increased C2 signature activity, we analyzed its relationship with additional clinical and laboratory parameters. The C2 signature showed a significant positive correlation with white blood cell count and C-reactive protein levels in both cohorts (**Fig. S14A**; s. **Tables S9 and S10** for correlation coefficients). Moreover, in the PREDICT cohort, patients with bacterial infection within the SDC and UDC groups displayed significantly higher C2 signature activity compared with patients without infection (**Fig. S14B**).

Finally, Kaplan–Meier analyses demonstrated that patients with high C2 signature activity at hospital admission had a significantly reduced ACLF-free probability and overall survival compared with patients with low C2 signature activity in both the PREDICT and ACLARA cohorts (**Fig. 6E**).

In summary, we demonstrated that the C2 signature can be detected in whole blood RNA-seq data from AD patients and, crucially, is associated with bacterial infection, progression to ACLF and mortality in two independent cohorts.

### Functional validation confirms impaired Complex IV activity in AD patients progressing to ACLF

To further validate the relevance of impaired energy metabolism in AD patients progressing to ACLF, we conducted respirometry analyses in an additional prospective, single-center cohort of 12 AD patients (**Fig. 1C, Table S11**). Patient recruitment took place at the center in Aachen, Germany, between 2021 and 2024, and the inclusion and exclusion criteria were identical to the single-cell study cohort (**Table S1**). After 90 days of follow-up, 8 patients were classified as SDC or UDC, and 4 as pre-ACLF (ACLF development within 28 days). Whole blood samples were collected at hospital admission, and classical monocytes were subsequently isolated. Mitochondrial function was assessed using Respirometry in Frozen Samples (RIFS) on a Seahorse Extracellular Flux Analyzer (Agilent), targeting the entire classical monocytes population.

Notably, despite analyzing the total CD14+ monocyte pool, we observed a progressive decline of the Complex IV activity, measured by the oxygen consumption rate (OCR), in pre-ACLF patients compared to SDC and UDC (**Fig. 7C**). In contrast, no significant differences were detected in the maximal respiratory capacity driven by complexes I and II (**Fig. 7A, B**). Notably, this decline in Complex IV function mirrors the transcriptional downregulation observed specifically in the C2 subset, which constitutes only a fraction of the classical monocyte population. The persistence of this signal at the whole-population level highlights the biological relevance of our findings and supports a model in which monocyte metabolic dysfunction contributes to AD progression and ACLF development.

**Fig. 7.**
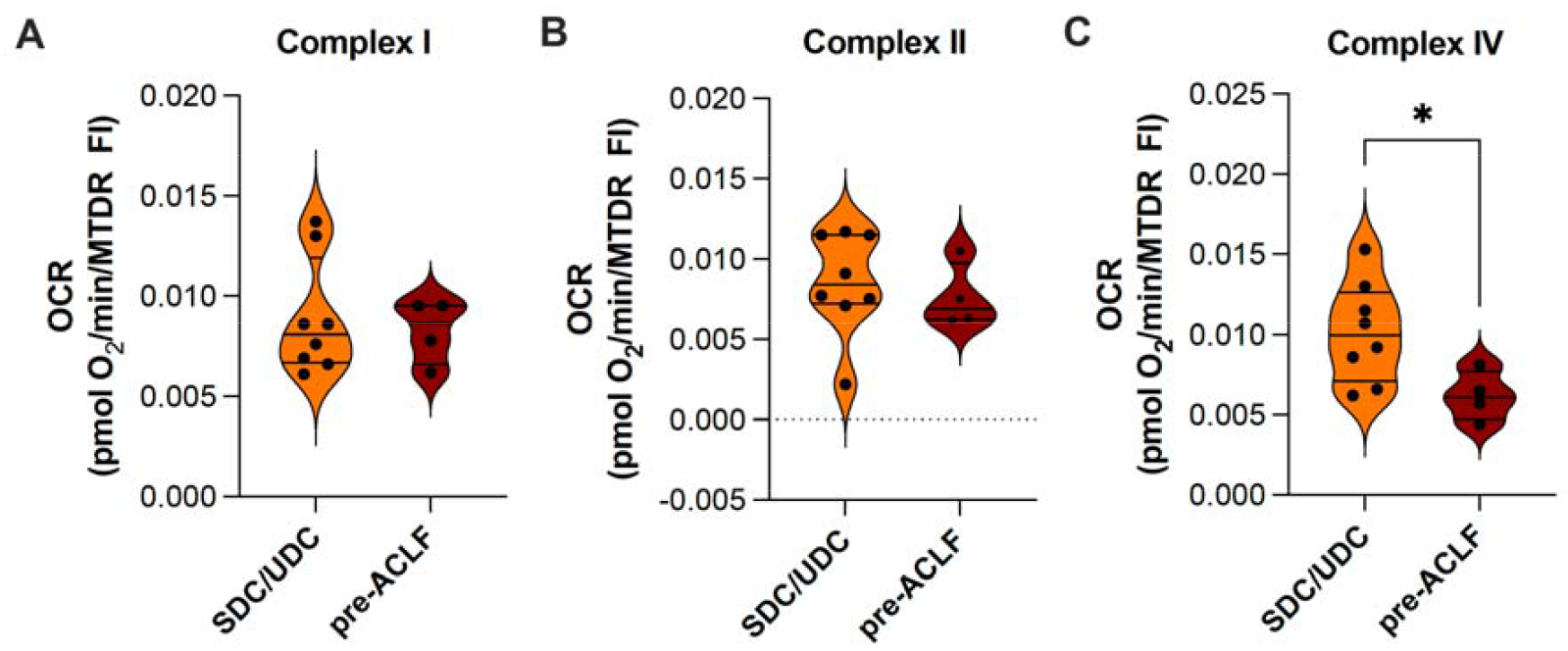
Functional validation confirms impaired Complex IV activity in an independent cohort of AD patients progressing to ACLF. Violin plots showing oxygen consumption rates (OCR) driven by respiratory chain complexes I **(A)**, II **(B)**, and IV **(C)** in CD14+ monocytes from patients with SDC/UDC and pre-ACLF. The OCR was measured by respirometry using a Seahorse Extracellular Flux Analyzer. Each point represents a single patient. Statistical significance was assessed using unpaired t-tests. * *p* < 0.05. *Abbreviations:* OCR, oxygen consumption rate; pre-ACLF, pre-acute-on-chronic liver failure; SDC, stable decompensated cirrhosis; UDC, unstable decompensated cirrhosis.

## DISCUSSION

In this prospective European multicenter study, we investigated the transcriptomic alterations in peripheral immune cells at single-cell resolution in patients with AD. We identified classical monocytes as the immune cell population with the most pronounced transcriptional changes during disease progression and defined a distinct monocyte subset, termed C2, whose expansion was associated with the subsequent development of ACLF. This subset was characterized by impaired cellular energy metabolism. Importantly, the transcriptional signature of C2 monocytes was associated with bacterial infection, ACLF development, and mortality in whole-blood RNA-seq data from two large independent cohorts of patients with AD.

Unlike previous case–control studies that focused on patients with established ACLF (16–18), our prospective design enabled the identification of immune alterations that preceded ACLF. While prior studies have reported enrichment of monocytic gene signatures in patients with manifest ACLF (15,16), our data demonstrate that these transcriptional alterations emerge earlier during AD, highlighting monocytes as key contributors to disease progression.

Subclustering of classical monocytes revealed three subsets (C0–C2) aligned along a pseudotime trajectory, with C2 selectively expanded in pre-ACLF patients (up to 10%) and correlated with a shorter latency to ACLF. The presence of C2 monocytes at low frequencies in the healthy controls and other AD groups suggests that this population exists under physiological conditions and expands with disease progression.

We next examined whether the C2 subset corresponds to previously described immunoregulatory monocyte populations, including mononuclear myeloid-derived suppressor cells (M-MDSCs) and endotoxin-tolerant monocytes. Although reduced CD14 and HLA-DR expression partially overlapped with features of M-MDSCs, several defining characteristics of this population were absent, arguing against a classical M-MDSC phenotype (20). Similarly, comparison of the C2 transcriptomic profile with established datasets of endotoxin-tolerant, i.e. sepsis-associated monocytes, revealed only limited overlap (21). Taken together, these findings indicate that C2 monocytes do not correspond to either M-MDSCs or endotoxin-tolerant monocytes, supporting the concept that C2 represents a previously undescribed and distinct monocyte state associated with AD progression and subsequent ACLF development.

To further characterize the C2 subpopulation, transcriptomic profiling was conducted using marker genes for monocyte differentiation and expansion during cirrhosis progression. Various genes encoding proteins related to inflammatory and immune responses, including interleukin-1 receptor-associated kinase 3 (*IRAK3*), hypoxia-inducible factor 1α (*HIF-1α*), and Toll-like receptor 2 (*TLR2*), were upregulated in the C2 subset. In contrast, genes associated with *suppressed* immune responses were increasingly expressed, including *MERTK*, which is associated with immunoregulatory monocytes in patients with ACLF (22) and *VCAN*, encoding versican, which was found to be associated with an immunosuppressive phenotype of monocytes in sepsis (23). Conversely, genes encoding pro-inflammatory S100 proteins (*S100A8, S100A9, S100A12)*, and key genes relevant to the bacterial immune response, such as *CD68* and *LYZ*, were downregulated in the C2 subset. These findings suggest that C2 monocytes exhibit an immunoregulatory phenotype marked by the upregulation of genes involved in immune modulation and inflammatory signaling, alongside downregulation of genes related to antibacterial defense. These alterations are consistent with early immune dysfunction and may contribute to an increased susceptibility to bacterial infection, a key precipitating event in ACLF. This concept was supported by whole-blood RNA-seq analyses in the PREDICT and ACLARA cohorts, in which the C2 signature correlated with disease severity and was enriched in patients with bacterial infections.

Pathway analyses identified suppression of cellular energy metabolism as the dominant feature of C2 monocytes. In contrast, immune-related pathways showed only modest changes. Functional validation using Seahorse respirometry in an independent AD cohort confirmed impaired Complex IV–dependent respiration in classical monocytes from pre-ACLF patients. Notably, this defect was detectable in the entire monocyte population, underscoring the biological relevance of C2-associated metabolic dysfunction.

Impaired cellular energy metabolism compromises key immune functions, including phagocytosis, cytokine production, and antigen presentation, and is associated with adverse outcomes in systemic inflammatory conditions such as sepsis (24). In the context of AD, bioenergetic failure in monocytes may limit effective immune responses and promote susceptibility to infection, thereby facilitating the progression to ACLF.

Our findings align with those of previous reports describing progressive metabolic dysfunction across the spectrum of cirrhosis, AD, and ACLF, likely driven by systemic inflammation (17,25–28). The identification of a distinct monocyte subset characterized by early bioenergetic impairment provides further support for the concept of inflammation-associated energetic exhaustion preceding ACLF development (14,29). Given that systemic inflammation in cirrhosis is partly driven by bacterial translocation, these findings may have implications for therapeutic strategies targeting inflammatory and metabolic pathways, including interventions aimed at modulating the gut barrier function and microbiota composition (30,31). Moreover, this subset could serve as a model for developing and testing therapeutic strategies aimed at restoring immune cell function and preventing ACLF progression.

This study had several limitations. Although study integrates single-cell, bulk transcriptomic, and functional data, additional mechanistic experiments are required to directly link monocyte bioenergetic failure to disease progression. Moreover, longitudinal single-cell profiling would further clarify the temporal dynamics of monocyte subset evolution during AD.

In summary, we identified a novel classical monocyte subset associated with the progression of AD to ACLF. This subset is characterized by immunoregulatory features and pronounced bioenergetic dysfunction, which may increase susceptibility to bacterial infections and contribute to disease progression. Targeting immune cell metabolic exhaustion may represent a promising strategy for preventing ACLF development in patients with cirrhosis.

## Data Availability

Single-cell data produced in the present study are available upon reasonable request to the authors. A submission to the European Genome-phenome Archive (EGA) is under progress.
Bulk RNA-seq data used in the present study are available upon reasonable request to the authors.

## Data and Code Availability

Extended details of the entire analysis pipeline and figure generation code are available at https://github.com/TranslationalBioinformaticsUnit/C2_ACLF_2025. The raw and processed sequencing data generated in this study have been deposited in the European Genome-phenome Archive (EGA) under the accession number <u>EGAS50000001127</u>.

## Acknowledgements

We thank all participating centers for their efforts in patient recruitment and fruitful collaboration. We thank the Genomics Facility of the Interdisciplinary Center for Clinical Research (IZKF) Aachen for sequencing experiments. Special thanks go to Jasmin Hübner and Anna Rudzinski for their extraordinary help in sequencing the samples.

## Authors’ contributions

Conceptualization: THW, RK, JC, PER, CT, and DGC

Supervised patient recruitment and sample collection and assessed clinical data: THW, MRP, MB, GZ, DB, MS, WG, RS, SV, AV, DC, and IG

Conducted experiments: THW, MRP, FS, AMA, JR and LS.

Data assessment, analysis, and visualization – lead: SPE.

Data assessment, analysis, and visualization – support: SPE, THW, DGC, NP, CLV and EH

Data interpretation: THW, SPE, MRP, LS, DGC, NP, and EH.

Writing – original draft: THW, SPE, NP, and DGC.

Writing – review and editing: EH, MRP, FS, JR, AK, RK, TB, PC, CA, NK, LS, JT, JC, PER, and CT.

Writing – final review: all authors.

## Financial support

This project has received funding from the European Union’s Horizon 2020 research and innovation program under grant agreement No 847949. This research was made possible through access to data generated by the PREDICT and ACLARA studies promoted and funded by the European Foundation for the Study of Chronic Liver Failure (EF-CLIF), a private, non-profit organization supported by unrestricted grants from Grifols and Genfit.

T.H.W. received financing from the START program of the RWTH Aachen University and from the Andrew K. Burroughs Short-Term Fellowship of the European Association for the Study of the Liver (EASL).

N.P.P. was funded by a Ramón y Cajal fellow (RYC2021-032197-I) from the MCIN/AEI/10.13039/501100011033 and European Union “NextGenerationEU”/PRTR and by a Juan

de la Cierva-formación fellow (FJC2019-042304-I) from the Spanish Ministry of Science and Innovation (MCIN).

T.B. was supported by the German Research Foundation (CRC1382 Project ID 403224013/B07).

The Genome Analysis Platform in CIC bioGUNE was supported by the Basque Department of Industry, Tourism and Trade (Etortek, Elkartek and Emaitek Programs), the Innovation Technology Department of Bizkaia County, CIBERehd Network, and Spanish MINECO the Severo Ochoa Excellence Accreditation (CEX2021-001136-S).

JC laboratory is supported by Ministerio de Ciencia e Innovación/Agencia Estatal Investigación MCIN/AEI (PID2022-138970OB-I00 10.13039/501100011033/FEDER, UE). CT was supported by the German Research Foundation grants TR285/10-2 and SFB1382, Project ID 403224013/A08.

PER research laboratory is supported by the Fondation pour la Recherche Médicale (FRM EQU202303016287), “Institut National de la Santé et de la Recherche Médicale” (ATIP AVENIR), the “Agence Nationale pour la Recherche” (ANR-18-CE14-0006-01, RHU QUID-NASH, ANR-18-IDEX-0001, ANR-22-CE14-0002) by « Émergence, Ville de Paris », by Fondation ARC and by France 2030 RHU LIVER-TRACK (ANR-23-RHUS-0014).

D.G.-C. research laboratory was supported by the King Abdullah University of Science and Technology.

