## Supplementary Material for "Single-cell profiling reveals monocyte mitochondrial dysfunction in patients with cirrhosis progressing to acute-on-chronic liver failure"

**Includes:**

- **Supplementary Methods.**
- **Supplementary Tables.**
- **Supplementary Figures.**

**Supplementary Methods**

### Sample collection and PBMC isolation

After study inclusion, peripheral blood was drawn from all participants from the single-cell study cohort, using BD vacutainer CPT tubes (BD Biosciences;(1)) for PBMC isolation and TempusTM tubes (Applied Biosystems) for whole blood assessment. After density gradient centrifugation at 1500 RCF for 20 minutes at 20°C PBMCs were aspirated and then cryopreserved using fetal calf serum with 10% DMSO. The cells were gradually frozen and stored at -80°C for several weeks until library preparation and sequencing or flow cytometry analysis, respectively.

### CITE-seq

#### Library preparation, sequencing, and demultiplexing

The isolated PBMCs were recovered by gently thawing and immediately diluting in pure FCS. Next, cells were centrifuged at 500 RCF for 7 minutes at 20°C, and the supernatant was removed. PBMCs were then resuspended in 4 ml 5% FCS in PBS and filtered through a 20 µm pluriStrainer**^®^** provided by pluriSelect (Leipzig, Germany) to obtain a single-cell solution. After the PBMCs were passed through a strainer, a second centrifugation was performed. The supernatant was removed, and cells were diluted in 100 µl 5% FCS in PBS. 5 µl Human TruStain FcX (BioLegend) was added and incubated for 10 minutes at 4°C.

A panel of 23 TotalSeq-C antibodies (BioLegend) was pooled, using 0.5 µg of each antibody (**Table S13**). Samples were incubated with the TotalSeq-C antibody pool for 30 min at 4°C. Cells were washed two times using 3 ml 5% FCS in PBS and rediluted in 1.5 ml of sorting buffer (5% FCS in PBS).

For the detection of dead cells, DAPI was added at a final concentration of 0.5 µg/ml. Up to 300,000 cells from each sample were sorted on a Sony Cell Sorter by life/dead sorting. Sorted viable cells were resuspended in 10 µl 1% BSA in PBS, counted, and ~20,000 cells in 38.7 µl were applied to single cell partitioning and barcoding *via* Chromium Controller (10x Genomics).

Each participant's sample was prepared into two distinct libraries: (I) 5′ Gene Expression Library (GEX), designed for scRNA-seq to capture transcriptome data; and (II) Surface Protein Library, which utilized TotalSeq-C barcodes to profile surface proteins, aligning with the CITE-seq methodology.

Cell Ranger (v6.01) was used for read alignment, filtering, barcode processing, and UMI counting with default settings (2). GRCh38-2020 was used as the transcriptome reference. Specifically, STAR was used in FASTQs alignment (3). Cell barcodes were then automatically determined based on the distribution of the UMI count.

#### Pre-processing

We used the R package *Seurat* v4.3.0 (4) to preprocess the gene-barcode matrix and the surface protein matrix of both patients and healthy controls and to perform further analysis. For each sample, genes not observed in at least three cells were dropped. Low-quality or damaged cells were excluded using a combination of a number of expressed genes (cells with <300 and >3500 genes) and mitochondrial transcript percentages (cells with >10% mitochondrial genes). Each sample dataset was normalized using the *‘SCTransform’* function (5) from the *Seurat* package and regression of mitochondrial RNA and cell cycle genes.

#### Integration, clustering, and cell type identification

The filtered and cleaned single-cell dataset underwent a process to identify highly variable genes and remove unwanted sources of variation. This was done by calculating and extracting a subset of features that show high cell-to-cell variation in the dataset using Seurat's ‘FindVariableFeatures’ function. Additionally, the ‘FeatureScatter’ plot from Seurat was applied, and we removed the detected outliers when plotting feature-feature relationships (6). To account for technical batch effects introduced because not all samples were sequenced at the same time, we applied Harmony with default parameters after dimensionality reduction (7). This correction improved integration across datasets while preserving biological variability. For each sample where the protein surface markers data were available, we selected the most variable protein subspace and applied feature-wise centered log-ratio transformation (CLR) (8). Data were visualized using UMAP. Data were scaled, and the dimensionality was reduced using PCA. To identify cell clusters, a shared nearest neighbor (SNN) graph was built using distances in PCA space (top 1 to 20 components) through Seurat's ‘FindNeighbors’ function. The Louvain algorithm was employed using Seurat’s ‘FindClusters’ function. For the global analysis including all cell types, we used a resolution parameter of 0.5, which provided clusters consistent with major immune cell lineages as confirmed by canonical marker expression (Fig. S2), surface protein markers (Fig. S3A, B), and Azimuth reference mapping (4). For the monocyte subset analysis, we applied clustering with a resolution of 0.15, which produced three distinct clusters that aligned with the pseudotime trajectory. Single-cell trajectory inference was performed using the ‘learn_graph’ function from the Monocle3 package with default parameters (9). Cells were subsequently ordered and visualized along pseudotime. This approach ensured that the chosen clustering resolution reflected biologically meaningful heterogeneity within the monocyte population.

#### Cluster Composition Analysis: changes in cell proportions between groups

Differences in cell type proportion between conditions were assessed by comparing the proportional differences in cell populations between AD patients and healthy controls for both gene expression and protein surface markers data, utilizing the *‘sc_utils’* function from *scProportionTest* package with default parameters (10). Most most prevalent cell subtypes (those representing at least 1% of the total cells) were investigated. Estimations of the p-value and the confidence interval for the magnitude of the difference are computed, respectively, using a permutation test and bootstrapping.

#### Differential expression analysis and pathway analysis

Seurat *‘FindAllMarkers’* function with the default Wilcoxon rank sum test based on the normalized data was used to identify the cluster markers used for cluster annotation. The ‘*FindMarkers’* function was employed to identify markers for each cell cluster to measure differential gene expression between conditions, using the default Wilcoxon rank sum test method. Genes detected in less than 50% of both evaluated groups were discarded. We considered differentially expressed genes (DEG) to be statistically significant if the adjusted p-value < 0.05 (Benjamini-Hochberg (BH)) and absolute Fold Change (FC) > 1.5. For the derivation of the C2 gene signature at the whole-blood level, genes identified at single-cell resolution were further filtered based on bulk RNA-seq data. Only genes showing a fold change >25% in bulk RNA-seq (log₂FC > 0.32) when comparing pre-ACLF patients with healthy controls, SDC, and UDC were retained for signature construction. ‘*VlnPlot’*, ‘*FeaturePlot*,’ and ‘*Dotplot’* functions were applied to visualize gene expression in each cell type or group.

Next, Gene Set Enrichment Analysis (GSEA) was performed using the R package *ClusterProfiler* (11), considering the comprehensive functions included in Gene Ontology (GO). Biological pathways in which genes appeared at a different frequency than expected were considered enriched. The results were visualized using default enrichment plots. Enrichment scores with p-value adjusted < 0.05 were considered significant. The differences in the pathways among patient groups were computed using the non-parametric Kruskal Wallis test.

#### Module scores for specific gene sets

We utilized cell module scores to quantify the extent of individual cell expression of specific predefined gene sets. For this purpose, we used the ‘*AddModuleScore’* function from *Seurat* with default settings. The lists of genes defining this module were prepared based on the intersection of upregulated DEGs resulting from various differential expression analyses (DEA). Differences in module scores were assessed using the non-parametric Kruskal Wallis test.

### Bulk RNA-seq

#### Library preparation, sequencing and demultiplexing

Bulk RNA-seq was performed in collaboration with the Genomics platform of the Center for Cooperative Research in Biosciences (CIC bioGUNE), Derio, Bizkaia, Spain. RNA was extracted from whole blood collected in Tempus^TM^ tubes from the single-cell study cohort using the Extraction Tempus™ Spin RNA Isolation Kit (Applied Biosystems). The extracted RNA was treated with DNase. RNA quality was assessed using Agilent RNA 6000 Nano and Pico Chips (Agilent Technologies). RNA concentration was assessed using RNA HS Assay Kit in a Qubit 2.0 fluorometer (Thermo Fisher Scientific).

Sequencing libraries were prepared using the NuGEN UniversalPlus mRNA-Seq with NuQuant kit (Tecan Life Sciences, Cat.#0520-24) and following the user guide (M01485v6.1andv7). The kit is intended to prepare sequencing libraries depleted of globin. Briefly, starting from 500 ng of total RNA, rRNA, and globin mRNA were depleted, and the remaining RNA was purified, fragmented, and primed for cDNA synthesis. cDNA first strand was synthesized with SuperScript-II Reverse Transcriptase (Thermo Fisher Scientific, Cat.#18064-014) for 10 min at 25°C, 15 min at 42°C, 15 min at 70°C and pause at 4°C. The second strand of cDNA was synthesized with Illumina reagents at 16°C for 1 hour. Then, A-tailing and adaptor ligation were performed. Finally, enrichment of libraries was achieved by PCR (30 s at 98°C; 15 cycles of 10 s at 98°C, 30 s at 60°C, 30 s at 72°C; 5min at 72°C and pause at 4°C).

Afterward, libraries were visualized on an Agilent 2100 Bioanalyzer using Agilent High Sensitivity DNA kit (Agilent Technologies, Cat. # 5067-4626), quantified using Qubit dsDNA HS DNA Kit (Thermo Fisher Scientific, Cat. # Q32854) and sequenced in a NovaSeq-6000 (Illumina Inc.) by at least 40 million paired-end 100nt reads.

Raw reads were demultiplexed via bclfastq2 Conversion Software v2.20 (Illumina) to convert base call (BCL) files into FastQ files. Quality control of FastQ files was performed using FastQC (Bioinformatics Babraham Institute) (12), and reads were trimmed for contaminating sequence adapters and poor-quality bases using Trimmomatic (13). Sequencing reads were aligned to the Human (GRCh38) genome with STAR (2.6.1) setting the parameters to default values (3). Uniquely aligned reads were assigned to GENCODE18 gene annotations using HTSeq v0.11 and quantified with htseq-counts run in stranded mode with default parameters (14).

#### Bulk RNA-seq downstream analysis

R v4.3.0 was employed in the downstream analysis of RNA-seq data. Genes exhibiting more than 1 count per million (CPM) in at least three samples were selected for the analysis. Utilizing the *limma-voom* workflow (15), RNA-seq count data was normalized, and variance estimation was performed. PCA was applied to the normalized data to assess sample similarity.

#### Module scores

A module score was calculated for each evaluated sample group using the Gene Set Variation Analysis (*GSVA*) R package v1.42.0 (16) in the single-cell paired bulk RNA-seq, as well as in the PREDICT and the ACLARA cohorts. This gene signature was calculated using previously selected genes to compute the module score at the single-cell level.

### Flow Cytometry Analysis

Flow cytometry of peripheral CD14^+^ monocytes was conducted. For this purpose, additional PBMC aliquots from the single-cell study cohort were used. First, PBMCs were thawed gently, as described above. Subsequently, CD14^+^ cells were enriched by magnetic cell separation using positive selection for human CD14 (Miltenyi Biotec, Bergisch Gladbach, Germany) for in vitro experiments.

For surface marker expression analysis, CD14^+^ peripheral monocytes were incubated with fluorochrome-conjugated primary antibodies at optimal dilutions at 4°C for 30 min in PBS containing 0.5% fetal calf serum and 2 mmol/L EDTA. Single-color staining, isotype-matched controls (IMCs), and fluorescence minus one (FMO) controls were applied. The antibodies used were CD14 (clone HCD14), CD16 (3G8), and human leukocyte antigen-DR (HLA-DR) (G46-6).

Cell viability was determined with the LIVE/DEAD™ Fixable Aqua Dead Cell Stain Kit from Thermo Fisher and IMC mouse IgG1 κ and IgG2a κ isotype controls from BD Pharmingen (BD Biosciences, Franklin Lakes, NJ, USA). Flow cytometry analysis was performed using a FACS Canto II flow cytometer and FlowJo (v10) software (BD Biosciences, Franklin Lakes, NJ, USA). Delta mean fluorescence intensity (ΔMFI) was calculated by subtracting the mean MFI count in the SDC group from single MFI counts in the four study groups. The Wilcoxon–Mann–Whitney test was used to test statistical differences within the four study groups.

*Validation cohorts*

*Validation studies* were conducted using whole blood bulk RNA-seq data from two prospective, multicenter European and Latin America cohorts, PREDICT and ACLARA (17,18). The inclusion and exclusion criteria of both cohorts are shown in **Tables S13 and S14**. Both cohorts collected various measures, including clinical, pharmacological, and outcome data from AD patients upon hospital admission and during follow-up visits (**Tables S6 and S7**). The follow-up period was 90 days for PREDICT and 28 days for ACLARA (**Fig S7 and S8**). 90 days after study inclusion, AD patients from the *PREDICT* cohort were categorized into SDC/UDC and pre-ACLF (18). *ACLARA* patients were classified either into AD or pre-ACLF. The diagnostic criteria of ACLF in both cohorts were according to the EF-CLIF definition (17).

The criteria for inclusion in our investigation were AD at hospital admission, availability of data on short-term outcomes (liver transplant, ACLF development, or death), clinical data, treatment, and availability of biological samples (buffy coat, tempus, serum, and plasma). Based on these criteria, 768 AD patients from PREDICT were chosen for analysis, with 738 samples passing quality control after sequencing. According to the definition of ACLF within the single-cell study cohort, only patients who developed ACLF within 28 days after hospital admission were included (n=61, **Fig. S9**). For the ACLARA cohort, the same criteria applied, with the added requirement that samples be available at the Barcelona EFCLIF biobank by the end of March 2021 due to COVID-19-related delays in shipping from South America. From ACLARA, 549 AD cirrhosis patients were selected, with 32 lacking buffy coat samples and 27 failing quality control after sequencing (**Fig. S10)**. In total, 689 patients from PREDICT and 521 patients from ACLARA were included to validate the single-cell results.

### PREDICT & ACLARA bulk RNA-seq

Whole blood RNA-seq from 1,210 patients with AD included in the PREDICT (n=689) (17) and the ACLARA (n=521) (18) studies were conducted according to the above-described bulk RNA-seq protocol.

A module score was calculated for each evaluated sample group, obtaining a gene signature score, as described in Material and Methods: Module scores. Pearson (parametric) and Spearman (non-parametric) correlation coefficients were computed to assess the relationships between the module score and clinical variables. The analysis of mean differences in the computed module scores among patient groups was assessed using the ANOVA test when more than two groups were compared and the Wilcoxon Signed-Rank test otherwise. Kaplan-Meier survival curves were used to assess progression-free survivaland ACLF-free survival across different subgroups in both the Predict and Aclara cohorts. Patients were stratified based on their scaled C2 values, with those having negative values categorized as "low" and those with positive values as "high."

*Respirometry analysis*

*Validation cohort and classical monocyte isolation*

To validate the single-cell results, we analyzed an independent cohort of 12 patients who were prospectively recruited to the University Hospital in Aachen, Germany, between 2021 and 20204 due to AD (**Table S15**). Inclusion/exclusion criteria, clinical data collection and whole blood sample acquisition were applied or performed respectively following the same procedure as for the single-cell study cohort (**Table S10**). After 90 days follow-up, patients were classified as SDC, UDC (“SDC/UDC”, n=8) or pre-ACLF (n=4) with ACLF development within 28 days.

Peripheral blood was drawn from all participants using SARSTEDT S-Monovette K3 EDTA. Mononuclear cells were isolated by density gradient centrifugation at 1500 RCF for 20 minutes at 20°C with Lymphocyte-H separation media (Cedarlane, Burlington, Ontario, Canada). CD14+ cells were enriched by magnetic cell separation using positive selection for human CD14 (Miltenyi Biotec, Bergisch Gladbach, Germany) and subsequently cryopreserved using fetal calf serum with 10% DMSO. Cells were gradually frozen and stored at -80°C for several weeks prior to respirometry analysis.

*Seahorse measurements and determination of cellular mitochondria content*

Respirometry analyses of classical monocyte samples was conducted according to the protocol for respiration measurement in previously frozen samples (“RIFS” (19)). Briefly, cells were thawed and washed in unsupplemented Mitochondrial Assay Solution (MAS; 70 mM Sucrose, 220 mM Mannitol, 5 mM KH_2_PO_4_, 5 mM MgCl_2_, 1 mM EGTA, 2 mM HEPES; pH 7.2). Cell counts were determined and adjusted to 12.5x10^6^/ml by resuspension in MAS. Next, cells underwent three freeze-thaw cycles with liquid nitrogen before use.

For Seahorse assay, 20 µL of the cell lysate with 250,000 cells/well loaded onto XFe96 microplates on the day of the assay. The samples were centrifuged at 2100×g for 10 min and stopped without a break. In a next step, samples were brought to a final volume of 150 µL with MAS supplemented with 100 µg/mL cytochrome c and 10 µg/mL alamethicin. Injections resulted in a final concentration of either 2 µM rotenone and 5 mM succinate or 1 mM NADH from Port A, 2 µM antimycin A and rotenone from Port B, 0.5 mM TMPD and 1 mM Asc from Port C, and 50 mM sodium azide from Port D.

OCR measurements included one baseline measurement before substrate injection (NADH or succinate/rotenone), two measurements after maximal substrate-driven respiration, two measurements post-antimycin A/rotenone injection, two measurements following TMPD/ascorbate addition, and two measurements after Complex IV inhibition with sodium azide. All measurements were performed in technical duplicates.

For normalization to mitochondria content, samples were stained with 500 nM MitoTracker Deep Red (MTDR) in PBS for 5 min in a black 96-well plate. After centrifugation at 2,100 xg for 5 minutes, wells were washed with PBS to remove the MTDR. Fluorescence intensity (FI) was measured from the bottom of the plate in a Tecan Spark 10 M Multimode Plate Reader using an excitation wavelength of 633 nm and an emission wavelength of 678 nm. OCR values were normalized to MTDR FI. Statistical differences between AD patients without (SDC/UDC) and with ACLF development (pre-ACLF) within 28 days were assessed using an unpaired t-test.

#### Statistical Methods

All analyses were performed in the R Computing Environment version 4.0.3 (20). Statistical methods are described in detail above in each subsection. In all statistical analyses, we considered *p*-value < 0.05 as statistically significant. In case an adjusted p-value for multiple testing is considered, it is described in the text.

*Study approval*

Written informed consent was obtained from the patient, her/his spouse, or the appointed legal guardian upon hospital admission. The study protocol was approved by the local ethics committee of each participating center (Paris, Frankfurt, Aachen, Bologna, Turin) and conducted in accordance with the Declaration of Helsinki.

*Data and material availability*

Researchers who provide a methodology sound proposal can apply for the data, as far as the proposal is in line with the research consented by the patients. These proposals should be requested through <https://www.clifresearch.com/decision/Home.aspx>. Data requestors will need to sign a data transfer agreement.

### List of Supplementary Materials

Fig S1 to S14

Tables S1 to S14

References included in the Supplementary Material

**Supplementary Tables**

***Table S1 Inclusion and exclusion criteria for single-cell study cohort.***

| Inclusion criterion | 1. The patient is admitted/referred to study center with AD of cirrhosis (ascites, overt encephalopathy, new onset of non-obstructive jaundice, GI-hemorrhage and/or bacterial infections), but without ACLF (as defined according to the CANONIC study) at study inclusion. |
| --- | --- |
| Exclusion criteria | 1. Pregnancy. 2. Age <18 years. 3. Patients with acute or subacute liver failure without underlying cirrhosis. 4. Patients who develop ACLF due to acute infection with Hepatitis E virus. 5. Patients with cirrhosis who develop decompensation in the postoperative period following partial hepatectomy. 6. Evidence of current malignancy except for non-melanocytic skin cancer and hepatocellular carcinoma within Milan criteria. 7. Presence or history of severe extra-hepatic diseases (e.g., chronic renal failure requiring hemodialysis, severe heart disease (NYHA > II), severe chronic pulmonary disease (GOLD > III), severe neurological and psychiatric disorders) 8. HIV-positive patients. 9. Previous liver or other transplantation. 10. Admission/referral of more than 72 hours before inclusion. 11. Patients who decline to participate or cannot provide prior written informed consent and when there is documented evidence that the patient has no legal surrogate decision maker, and it appears unlikely that the patient will regain consciousness or sufficient ability to provide delayed informed consent. 12. Physician’s denial (e.g. the investigator considers that the patient will   not follow the protocol scheduled). |

***Table S2 Visit and sampling schedule of single-cell study cohort.***

|  | **V1**  **Hospital admission/**  **Inclusion** | **A1 / A2**  **ACLF present or readmission** | **V2**  **Discharge** | **D1 / D2 / D3**  **Discharge: Recovery/death/OLT** | **F1 / F2 / F3 / F4 alive/dead/OLT**  **90d FU** |
| --- | --- | --- | --- | --- | --- |
| **Informed consents** | X | - | - | - | - |
| **Medical history**  Clinical data  Etiology of cirrhosis  Previous decompensations | X  X  X | -  -  - | -  -  - | -  -  - | -  -  - |
| **Clinical data**  AD score  ACLF score  Laboratory  Treatment | X  *(X, if ACLF present)*  X  X | X  *(X, if ACLF present)*  X  X | -  -  X  X | -  (X, *if ACLF cause of death*)  X  X | -  (X, *if ACLF cause of death*)  X  - |
| **Blood collection**  Serum  PBMC (Vacut.)  RNA (Tempus) | X  X  X | -  -  - | -  -  - | -  -  - | -  -  - |

*Abbreviations:* V, visit; A, admission; D, discharge; F, follow-up visit; OLT, orthotopic liver transplant; AD, acute decompensation; ACLF, acute-on-chronic liver failure; PBMC, peripheral blood mononuclear cells; Vacut., BD Vacutainer tubes.

***Table S3. Characteristics of single-cell study cohort patients with ACLF development within 28 days.***

| **Study variable** | **Pre-ACLF 1** | **Pre-ACLF 2** | **Pre-ACLF 3** | **Pre-ACLF 4** |
| --- | --- | --- | --- | --- |
| **Age** | 70 | 60 | 79 | 74 |
| **Sex** | male | male | female | female |
| **Previous clinical course** | 3 months history of cirrhosis, no previous decompensation, no ACLF | 14 years history of cirrhosis, >2 previous decompensations, no ACLF | 9 years history of cirrhosis, 1-2 previous decompensations, no ACLF | 2 years history of cirrhosis, 1-2 previous decompensations, no ACLF |
| **CLIF-C AD score**  **at admission** | 74 | 66 | 55 | 56 |
| **MELD score at admission** | 27 | 17 | 19 | 21 |
| **Days until ACLF development** | 6 | 4 | 10 | 5 |
| **ACLF grade** | 3 | 1 | 1 | 1 |
| **Type of decompensation** | ascites, alcoholic hepatitis | ascites, HE III°, SBP | ascites | ascites, SBP |
| **Organ failure** | hepatic + renal + coagulation + cerebral | renal | renal (+HE I°) | renal |
| **90d outcome** | non-survivor | survivor (liver transplant) | Transplant-free survivor | Transplant-free survivor |
| **Reason for death** | ACLF | - | - | - |

*Abbreviations:* ACLF: acute-on-chronic liver failure; SBP: spontaneous bacterial peritonitis; HE: hepatic encephalopathy.

***Table S4. Detailed baseline characteristics of single-cell study cohort.***

|  | **HC (n=4)** | **SDC**  **(n=6)** | **UDC**  **(n=6)** | **Pre-ACLF**  **(n=4)** | ***p*-value** |
| --- | --- | --- | --- | --- | --- |
| **Characteristics** | | | | | |
| Age, median (range) | 56 (52 - 64) | 60 (54 - 81) | 57 (51- 60) | 74 (60 - 79) | 0.063 |
| Female Sex, yes, n (%) | 2 (50) | 2 (33) | 0 | 2 (50) | 0.248 |
| Previous decompensations in the last 2 years, yes, n (%) | - | 0 | 4 (67) | 3 (75) | **0.023** |
| Hepatocellular carcinoma, yes, n (%) | - | 0 | 0 | 1 (25) | 0.202 |
| Alcohol consumption, active, n (%) | - | 1 (17) | 2 (33) | 1 (25) | 0.484 |
| **Cause of cirrhosis, n (%)** | | | | | |
| Alcohol-related | - | 1 (17) | 3 (50) | 2 (50) | 0.606 |
| Viral (HBV/HCV) | - | 1 (17) | 1 (17) | 0 | 0.259 |
| Alcohol-related + MASLD | - | 1 (17) | 1 (17) | 0 | 0.259 |
| MASLD | - | 2 (50) | 1 (17) | 1 (25) | 0.801 |
| cryptogenic | - | 0 | 0 | 1 (25) | 0.202 |
| other | - | 1 (17) | 0 | 0 | 0.411 |
| **Data at admission, n (%)** | | | | | |
| Ascites | - | 5 (83) | 5 (83) | 4 (100) | 0.683 |
| SBP | - | 0 | 0 | 2 (50) | **0.032** |
| Other bacterial infection | - | 2 (33) | 0 | 0 | 0.149 |
| Variceal bleeding | - | 0 | 1 (17) | 0 | 0.411 |
| HE | - | 1 (17) | 2 (33) | 1 (25) | 0.801 |
| Alcoholic hepatitis |  | 1 (17) | 0 | 1 (25) | 0.467 |
| Acute kidney injury | - | 0 | 1 (17) | 0 | 0.411 |
| **Laboratory parameters** | | | | | |
| Bilirubin, total [mg/dl], median, (range) | - | 2.31 (1.3-3.4) | 2.14 (1.37-4.60) | 3.91 (2.61-5.85) | 0.217 |
| AST [U/l], median, (range) |  | 44 (28-64) | 61 (35-361) | 54 (34-125) | 0.294 |
| ALT [U/l], median, (range) |  | 33 (11-38) | 32 (17-76) | 36 (18-57) | 0.709 |
| INR, median, (range) |  | 1.38 (1.2-1.75) | 1.46 (1.31-1.77) | 1.69 (1.26-2.95) | 0.516 |
| Creatinine [mg/dl], median, (range) |  | 0.72 (0.54-0.92) | 0.8 (0.6-1.97) | 1.36 (1.17-1.62) | **0.012** |
| Platelets [x10^3^/L], median (range) |  | 79 (39-176) | 75 (41-130) | 81 (37-106) | 0.972 |
| WBC [x10^9^/L], median (range) | 4.8 (4.0-6.3) | 4.1 (2.1-5.7) | 5.4 (2.9-7.5) | 5.5 (3.4-9.4) | 0.476 |
| CRP [mg/L], median (range) | - | 16.25 (2.22-76) | 13.7 (1.78-45.2) | 8.55 (1.23-37.1) | 0.567 |
| **Severity scores** | | | | | |
| Child-Pugh, median, range | - | 7.5 (5 - 12) | 10 (7 - 11) | 11 (8 - 13) | 0.126 |
| MELD, median, range | - | 12 (8 - 18) | 13 (12 - 23) | 20 (17 - 27) | **0.033** |
| CLIF-C AD, median, range | - | 47 (41 - 49) | 47.5 (41 - 59) | 61 (55 - 74) | **0.025** |
| **Treatment at blood draw** | | | | | |
| Diuretics, yes, n (%) | - | 3 (50) | 4 (67) | 3 (75) | 0.701 |
| Antibiotics, yes, n (%) | - | 1 (17) | 1 (17) | 1 (25) | 0.934 |
| Non-selective ß-blockers, yes, n (%) | - | 1 (17) | 2 (33) | 3 (75) | 0.169 |
| Albumin, yes, n (%) | - | 1 (17) | 0 | 1 (25) | 0.467 |
| HE treatment, yes, n (%) | - | 1 (17) | 3 (50) | 3 (75) | 0.176 |
| **Follow-up and Outcome** | | | | | |
| 90-day mortality, yes, n (%) | - | 0 | 0 | 1 (25) | 0.229 |
| 90-day OLT, yes, n (%) | - | 0 | 0 | 1 (25) | 0.229 |

HE treatment includes lactulose and rifaximin. Baseline characteristics are depicted as frequencies or median (IQR). Values of *p*-value are based on Wilcoxon–Mann–Whitney test for continuous and chi-square test for categorical variables. *Abbreviations:* SDC, stable decompensated cirrhosis; UDC, unstable decompensated cirrhosis; pre-ACLF, pre-acute-on-chronic liver failure; HBV: Hepatitis B virus; HCV: Hepatitis C virus; MASLD: metabolic-dysfunction associated steatotic liver disease; SBP: spontaneous bacterial peritonitis; HE: hepatic encephalopathy; AST, aspartate aminotransferase; ALT, alanine aminotransferase; WBC: white blood cell count; CRP: C-reactive protein; OLT: orthotopic liver transplantation.

***Table S5. Detailed overview on readmission and outcome of single-cell study cohort patients.***

| **Study ID** | **Readmission** | **If yes, when** | **ACLF development** | **If yes, when** | **90d outcome** | **If death cause** |
| --- | --- | --- | --- | --- | --- | --- |
| **SDC 1** | No | - | No | - | Transplant-free survivor | - |
| **SDC 2** | No | - | No | - | Transplant-free survivor | - |
| **SDC 3** | No | - | No | - | Transplant-free survivor | - |
| **SDC 4** | No | - | No | - | Transplant-free survivor | - |
| **SDC 5** | No | - | No | - | Transplant-free survivor | - |
| **SDC 6** | No | - | No | - | Transplant-free survivor | - |
| **UDC 1** | Yes | Day 56 | No | - | Transplant-free survivor | - |
| **UDC 2** | Yes | Day 18 | No | - | Transplant-free survivor | - |
| **UDC 3** | Yes | Day 49 | No | - | Transplant-free survivor | - |
| **UDC 4** | Yes | Day 43 | No | - | Transplant-free survivor | - |
| **UDC 5** | Yes | Day 76 | No | - | Transplant-free survivor | - |
| **UDC 6** | Yes | Day 74 | no | - | Transplant-free survivor | - |
| **Pre-ACLF 1** | -- | - | Yes | Day 6 | Non-survivor | ACLF |
| **Pre-ACLF 2** | - | - | Yes | Day 4 | Transplant | - |
| **Pre-ACLF 3** | - | - | Yes | Day 10 | Transplant-free survivor | - |
| **Pre-ACLF 4** | - | - | yes | Day 5 | Transplant-free survivor | - |

*Abbreviations:* SDC, stable decompensated cirrhosis; UDC, unstable decompensated cirrhosis; pre-ACLF, pre-acute-on-chronic liver failure.

***Table S6 GSEA GO biological pathways.***

| ID | Description | Set Size | Enrichment Score | NES | p-value | p.adjust |
| --- | --- | --- | --- | --- | --- | --- |
| GO:0006119 | oxidative phosphorylation | 50 | -0.7592 | -3.4230 | <0.0001 | **<0.0001** |
| GO:0009060 | aerobic respiration | 53 | -0.7383 | -3.3922 | <0.0001 | **<0.0001** |
| GO:0045333 | cellular respiration | 56 | -0.7247 | -3.3812 | <0.0001 | **<0.0001** |
| GO:0042773 | ATP synthesis coupled electron transport | 33 | -0.7908 | -3.2414 | <0.0001 | **<0.0001** |
| GO:0042775 | mitochondrial ATP synthesis coupled electron transport | 33 | -0.7908 | -3.2414 | <0.0001 | **<0.0001** |
| GO:0015980 | energy derivation by oxidation of organic compounds | 62 | -0.6753 | -3.2405 | <0.0001 | **<0.0001** |
| GO:0019646 | aerobic electron transport chain | 31 | -0.7883 | -3.1919 | <0.0001 | **<0.0001** |
| GO:0022904 | respiratory electron transport chain | 35 | -0.7640 | -3.1827 | <0.0001 | **<0.0001** |
| GO:0022900 | electron transport chain | 40 | -0.7401 | -3.1814 | <0.0001 | **<0.0001** |
| GO:0006091 | generation of precursor metabolites and energy | 89 | -0.5957 | -3.0521 | <0.0001 | **<0.0001** |
| GO:0015986 | proton motive force-driven ATP synthesis | 33 | -0.7392 | -3.0297 | <0.0001 | **<0.0001** |
| GO:1902600 | proton transmembrane transport | 38 | -0.7146 | -3.0230 | <0.0001 | **<0.0001** |
| GO:0009201 | ribonucleoside triphosphate biosynthetic process | 40 | -0.6891 | -2.9624 | <0.0001 | **<0.0001** |
| GO:0009206 | purine ribonucleoside triphosphate biosynthetic process | 40 | -0.6891 | -2.9624 | <0.0001 | **<0.0001** |
| GO:0042776 | proton motive force-driven mitochondrial ATP synthesis | 30 | -0.7374 | -2.9565 | <0.0001 | **<0.0001** |
| GO:0006754 | ATP biosynthetic process | 39 | -0.6867 | -2.9303 | <0.0001 | **<0.0001** |
| GO:0009152 | purine ribonucleotide biosynthetic process | 44 | -0.6584 | -2.8912 | <0.0001 | **<0.0001** |
| GO:0009142 | nucleoside triphosphate biosynthetic process | 41 | -0.6691 | -2.8807 | <0.0001 | **<0.0001** |
| GO:0009145 | purine nucleoside triphosphate biosynthetic process | 41 | -0.6691 | -2.8807 | <0.0001 | **<0.0001** |
| GO:0046390 | ribose phosphate biosynthetic process | 46 | -0.6521 | -2.8743 | <0.0001 | **<0.0001** |
| GO:0050729 | positive regulation of inflammatory response | 24 | -0.4506 | -1.7331 | 0.01 | 0.18 |
| GO:0042742 | defense response to bacterium | 34 | -0.3542 | -1.4649 | 0.049 | 0.36 |
| GO:0002700 | regulation of production of molecular mediator of immune response | 19 | -0.4029 | -1.4505 | 0.09 | 0.47 |
| GO:0002367 | cytokine production involved in immune response | 16 | -0.4205 | -1.4344 | 0.0892 | 0.47 |
| GO:0009617 | response to bacterium | 78 | -0.2785 | -1.3958 | 0.06 | 0.42 |
| GO:0002577 | regulation of antigen processing and presentation | 7 | -0.5361 | -1.3674 | 0.14 | 0.55 |
| GO:0002526 | acute inflammatory response | 10 | -0.4627 | -1.3368 | 0.16 | 0.58 |
| GO:0002821 | positive regulation of adaptive immune response | 19 | -0.3613 | -1.3009 | 0.18 | 0.61 |
| GO:0045087 | innate immune response | 122 | -0.2123 | -1.1378 | 0.25 | 0.69 |
| GO:0002237 | response to molecule of bacterial origin | 48 | -0.2470 | -1.0999 | 0.35 | 0.80 |

Results of the GO enrichment analysis were conducted on the C0 vs. the C2 cluster. The global top 20 pathways and the top 10 immune suppression and systemic inflammation-related pathways are displayed. The pathways in red are displayed in Figure 4. *Abbreviations:* NES, Normalized enrichment score.

***Table S7 Baseline characteristics of included patients from the PREDICT cohort.***

|  | **SDC (n=449)** | **UDC (n=179)** | **Pre-ACLF (n=61)** |
| --- | --- | --- | --- |
| **Characteristics** | | | |
| Age, median (range) | 58 (21-87) | 61 (33-87) | 60 (39-83) |
| Female Sex, yes, n (%) | 142 (31.6) | 56 (31.3) | 19 (31.2) |
| **Cause of cirrhosis, n (%)** | | | |
| Alcohol-related | 280 (63.4) | 101 (56.4) | 28 (45.9) |
| Viral (HBV/HCV) | 47 (10.5) | 21 (11.7) | 9 (14.8) |
| Alcohol-related + NASH | 37 (8.2) | 10 (5.6) | 9 (14.8) |
| NASH | 36 (8.0) | 19 (10.6) | 5 (8.2) |
| cryptogenic | 20 (4.4) | 11 (6.2) | 3 (4.9) |
| other | 29 (6.5) | 17 (9.5) | 7 (11.5) |
| **Main reason for admission, n (%)** | | | |
| Ascites | 189 (42.1) | 83 (46.4) | 23 (37.7) |
| HE | 65 (14.5) | 30 (16.8) | 9 (14.8) |
| GIB | 77 (17.2) | 27 (15.1) | 5 (8.2) |
| SBP | 13 (2.9) | 2 (1.1) | 4 (6.6) |
| Other | 48 (10.7) | 18 (10.1) | 6 (9.8) |
| **Laboratory parameters** | | | |
| Bilirubin, total [mg/dl], median, (range) | 2.26 (0.1-32.5) | 2.63 (0.25-32.5) | 4.20 (0.54-30.40) |
| AST [U/l], median, (range) | 50 (15-900) | 54 (13-431) | 56 (12-717) |
| ALT [U/l], median, (range) | 26.33 (6-695) | 29.34 (6.59-131) | 31 (7-334) |
| INR, median, (range) | 1.4 (0.87-9.99) | 1.43 (0.95-3.48) | 1.67 (1.2-3.01) |
| Creatinine [mg/dl], median, (range) | 0.85 (0.3-1.98) | 0.92 (0.35-1.99) | 1.10 (0.40-1.96) |
| Platelets [x10^3^/L], median (range) | 96 (8-516) | 97 (12-573) | 97 (11-283) |
| WBC [x10^9^/L], median (range) | 5.68 (0.79-26.25) | 5.90 (1.34-19.52) | 6.39 (2.6-27.76) |
| CRP [mg/L], median (range) | 13.3 (0.3-312) | 14 (0.3-172.7) | 21.01 (0.77-154.7) |
| **Severity scores** | | | |
| Child-Pugh, median, range | 9 (5-13) | 9 (5-13) | 11 (5-14) |
| MELD, median, range | 15 (6-40) | 16 (6-29) | 21 (9-29) |
| CLIF-C AD, median, range | 50 (31-86) | 53 (33-70) | 58 (42-76) |
| **Outcome** | | | |
| 90-day mortality, yes, n (%) | 0 (0) | 33 (18.44) | 37 (60.77) |
| 90-day OLT, yes, n (%) | 30 (6.7) | 11 (6.2) | 1 (1.6) |

Baseline characteristics are depicted as frequencies or median (IQR). *Abbreviations:* SDC, stable decompensated cirrhosis; UDC, unstable decompensated cirrhosis; pre-ACLF, pre-acute-on-chronic liver failure; HBV: Hepatitis B virus; HCV: Hepatitis C virus; NASH: non-alcoholic steatohepatitis; SBP: spontaneous bacterial peritonitis; HE: hepatic encephalopathy; GIB, gastrointestinal bleeding; AST, aspartate aminotransferase; ALT, alanine aminotransferase; WBC: white blood cell count; CRP: C-reactive protein; OLT: orthotopic liver transplantation.

***Table S8 Baseline characteristics of included patients from the ACLARA cohort.***

|  | **No ACLF development**  **(n=456)** | **ACLF development**  **(n=65)** |
| --- | --- | --- |
| **Characteristics** | | |
| Age, median (range) | 59 (18-96) | 60 (36-79) |
| Female Sex, yes, n (%) | 161 (35.3) | 24 (36.9) |
| **Cause of cirrhosis, n (%)** | | |
| Alcohol-related | 157 (34.4) | 30 (46.1) |
| Viral (HBV/HCV) | 63 (13.8) | 11 (16.9) |
| Alcohol-related + NASH | 26 (5.7) | 3 (4.6) |
| NASH | 77 (16.9) | 12 (18.5) |
| cryptogenic | 83 (18.2) | 5 (7.7) |
| other | 50 (11) | 4 (6.2) |
| **Main reason for admission, n (%)** | | |
| Ascites | 109 (23.9) | 15 (23.1) |
| HE | 78 (17.1) | 18 (27.7) |
| GIB | 147 (32.2) | 11 (16.9) |
| SBP | 41 (9) | 7 (10.8) |
| Other | 63 (13.8) | 13 (20) |
| **Laboratory parameters** | | |
| Bilirubin, total [mg/dl], median, (range) | 1.94 (0.30-26.15) | 2.75 (0.44-31.63) |
| AST [U/l], median, (range) | 40 (10-1096) | 54 (10-277) |
| ALT [U/l], median, (range) | 30 (5-578) | 30 (8-339) |
| INR, median, (range) | 1.48 (0.90-4.27) | 1.58 (1-3.16) |
| Creatinine [mg/dl], median, (range) | 0.9 (0.3-1.94) | 1.13 (0.39-1.92) |
| Platelets [x10^3^/L], median (range) | 88 (10-504) | 78 (4-325) |
| WBC [x10^9^/L], median (range) | 5.25 (0.85-33.70) | 5.54 (1.10-23.40) |
| CRP [mg/L], median (range) | 22.60 (0.66-209.3) | 30.4 (1.9-262.2) |
| **Severity scores** | | |
| Child-Pugh, median, range | 8 (5-13) | 10 (5-13) |
| MELD, median, range | 14 (6-31) | 19 (8-31) |
| CLIF-C AD, median, range | 51 (24-75) | 56 (35-72) |
| **Outcome** | | |
| 28-day mortality, yes, n (%) | 77 (16.9) | 44 (67.7) |
| 28-day OLT, yes, n (%) | 30 (6.6) | 3 (4.6) |

Baseline characteristics are depicted as frequencies or median (IQR). *Abbreviations:* SDC, stable decompensated cirrhosis; UDC, unstable decompensated cirrhosis; pre-ACLF, pre-acute-on-chronic liver failure; HBV: Hepatitis B virus; HCV: Hepatitis C virus; NASH: non-alcoholic steatohepatitis; SBP: spontaneous bacterial peritonitis; HE: hepatic encephalopathy; GIB, gastrointestinal bleeding; AST, aspartate aminotransferase; ALT, alanine aminotransferase; WBC: white blood cell count; CRP: C-reactive protein; OLT: orthotopic liver transplantation.

***Table S9 Detailed correlation analysis of C2 gene signature and baseline characteristics of included patients from the PREDICT cohort.***

|  | **Correlation value** | **p-value** |
| --- | --- | --- |
| **Age** | 0.00 | 0.95 |
| **Cause of cirrhosis** | | |
| Alcohol-related | -0.06 | 0.09 |
| Viral (HBV/HCV) | -0.03 | 0.51 |
| Alcohol-related + NASH | 0.01 | 0.75 |
| NASH | 0.07 | 0.06 |
| Cryptogenic | 0.05 | 0.20 |
| Other | 0.02 | 0.61 |
| **Main reason for hospital admission** | | |
| Ascites | -0.09 | **<0.05** |
| HE | -0.12 | **<0.001** |
| GIB | 0.00 | 0.91 |
| SBP | 0.04 | 0.28 |
| Other | 0.19 | **<0.001** |
| **Laboratory variables** | | |
| Bilirubin | 0.19 | **<0.001** |
| AST | 0.10 | **<0.05** |
| ALT | 0.09 | **<0.05** |
| INR | 0.01 | 0.72 |
| Creatinine | 0.08 | **<0.05** |
| Platelets | 0.09 | **<0.05** |
| WBC | 0.37 | **<0.001** |
| CRP | 0.30 | **<0.001** |
| **Severity scores** | | |
| CHILD | 0.11 | **<0.001** |
| MELD | 0.17 | **<0.001** |
| CLIF-C AD | 0.34 | **<0.001** |

*Abbreviations:* HBV: Hepatitis B virus; HCV: Hepatitis C virus; NASH: non-alcoholic steatohepatitis; SBP: spontaneous bacterial peritonitis; HE: hepatic encephalopathy; GIB, gastrointestinal bleeding; AST, aspartate aminotransferase; ALT, alanine aminotransferase; WBC: white blood cell count; CRP: C-reactive protein.

***Table S10 Detailed correlation analysis of C2 gene signature and baseline characteristics of included patients from the ACLARA cohort.***

|  | **Correlation value** | **p-value** |
| --- | --- | --- |
| **Age** | 0.13 | **<0.001** |
| **Cause of cirrhosis** | | |
| Alcohol-related | -0.08 | 0.08 |
| Viral (HBV/HCV) | -0.08 | 0.07 |
| Alcohol-related + NASH | 0.06 | 0.17 |
| NASH | 0.00 | 0.93 |
| Cryptogenic | 0.11 | **<0.05** |
| Other | 0.03 | 0.55 |
| **Main reason for hospital admission** | | |
| Ascites | -0.12 | **<0.05** |
| HE | 0.04 | 0.43 |
| GIB | 0.04 | 0.38 |
| SBP | 0.07 | 0.09 |
| Other | 0.03 | 0.56 |
| **Laboratory variables** | | |
| Bilirubin | 0.09 | **0.03** |
| AST | -0.02 | 0.73 |
| ALT | 0.01 | 0.86 |
| INR | 0.07 | 0.10 |
| Creatinine | 0.11 | <0.05 |
| Platelets | 0.04 | 0.31 |
| WBC | 0.30 | **<0.001** |
| CRP | 0.28 | **<0.001** |
| **Severity scores** | | |
| CHILD | 0.07 | 0.12 |
| MELD | 0.13 | **<0.001** |
| CLIF-C AD | 0.32 | **<0.001** |

*Abbreviations:* HBV: Hepatitis B virus; HCV: Hepatitis C virus; NASH: non-alcoholic steatohepatitis; SBP: spontaneous bacterial peritonitis; HE: hepatic encephalopathy; GIB, gastrointestinal bleeding; AST, aspartate aminotransferase; ALT, alanine aminotransferase; WBC: white blood cell count; CRP: C-reactive protein.

***Table S11 Baseline characteristics of validation cohort.***

|  | **SDC**  **(n=3)** | **UDC**  **(n=5)** | **pre-ACLF**  **(n=4)** |
| --- | --- | --- | --- |
| **Characteristics** | | | |
| Age, median (range) | 58 (45 - 66) | 56 (49- 68) | 54 (50 - 66) |
| Female Sex, yes, n (%) | 0 | 0 | 1 (25) |
| Previous decompensations in the last 2 years, yes, n (%) | 1 (33) | 4 (80) | 4 (100) |
| Hepatocellular carcinoma, yes, n (%) | 0 | 0 | 0 |
| Alcohol consumption, active, n (%) | 3 (100) | 5 (100) | 3 (75) |
| **Cause of cirrhosis, n (%)** | | | |
| Alcohol-related | 3 (100) | 4 (80) | 3 (75) |
| MASLD | 0 | 1 (20) | 1 (25) |
| **Data at admission, n (%)** | | | |
| Ascites | 3 (100) | 5 (100) | 4 (100) |
| SBP | 0 | 1 (20) | 2 (50) |
| Other bacterial infection | 1 (33) | 0 | 2 (50) |
| Variceal bleeding | 0 | 1 (20) | 0 |
| HE | 1 (33) | 2 (40) | 3 (75) |
| Alcoholic hepatitis | 0 | 0 | 1 (25) |
| Acute kidney injury | 0 | 1 (20) | 2 (50) |
| **Laboratory parameters** | | | |
| Bilirubin, total [mg/dl], median, (range) | 0.98 (0.71-2.34) | 0.81 (0.72-1.48) | 3.81 (3.22-4.68) |
| AST [U/l], median, (range) | 32 (22-65) | 37 (27-70) | 113 (89-125) |
| ALT [U/l], median, (range) | 12 (10-31) | 40 (16-64) | 80 (21-94) |
| INR, median, (range) | 1.19 (1.04-1.24) | 1.47 (1.18-1.61) | 1.70 (1.02-2.31) |
| Creatinine [mg/dl], median, (range) | 1.01 (0.9-1.15) | 1.49 (0.66-1.74) | 1.23 (1.11-1.59) |
| Platelets [x10^3^/L], median (range) | 90 (78-177) | 139 (112-181) | 104 (90-226) |
| WBC [x10^9^/L], median (range) | 6.2 (2.8-7.9) | 7.2 (6.0-12.9) | 10.0 (3.4-24.4) |
| CRP [mg/L], median (range) | 8.4 (7.9-19.1) | 10.25 (6-26.8) | 28.3 (8.5-96.3) |
| **Severity scores** | | | |
| Child-Pugh, median, range | 6 (6 - 8) | 10 (7 - 11) | 12 (10 - 12) |
| MELD, median, range | 11 (7 - 13) | 17.5 (14 - 20) | 27 (15 - 30) |
| CLIF-C AD, median, range | 51 (40 - 53) | 56 (54 - 67) | 63 (49 - 80) |
| **Treatment at blood draw** | | | |
| Diuretics, yes, n | 3 (100) | 4 (80) | 4 (100) |
| Antibiotics, yes, n | 1 (33) | 1 (20) | 1 (25) |
| Non-selective ß-blockers, yes, n | 1 (33) | 2 (40) | 4 (100) |
| Albumin, yes, n | 1 (33) | 0 | 1 (25) |
| HE treatment, yes, n | 1 (33) | 3 (60) | 3 (75) |
| **Follow-up and Outcome** | | | |
| 90-day mortality, yes, n | 0 | 0 | 4 (100) |

HE treatment includes lactulose and rifaximin. Baseline characteristics are depicted as frequencies or median (IQR). Values of *p*-value are based on Wilcoxon–Mann–Whitney test for continuous and chi-square test for categorical variables. *Abbreviations:* SDC, stable decompensated cirrhosis; UDC, unstable decompensated cirrhosis; pre-ACLF, pre-acute-on-chronic liver failure; HBV, Hepatitis B virus; HCV, Hepatitis C virus; MASLD, metabolic-dysfunction associated steatotic liver disease; SBP, spontaneous bacterial peritonitis; HE, hepatic encephalopathy; AST, aspartate aminotransferase; ALT, alanine aminotransferase; WBC, white blood cell count; CRP, C-reactive protein.

***Table S12 CITE-seq antibody panel.***

| Antibody | Clone | Source | Catalog number |
| --- | --- | --- | --- |
| TotalSeq™-C0048 anti-human CD45 Antibody | 2D1 | BioLegend | 368545 |
| TotalSeq™-C0034 anti-human CD3 Antibody | UCHT1 | BioLegend | 300479 |
| TotalSeq™-C0072 anti-human CD4 Antibody | RPA-T4 | BioLegend | 300567 |
| TotalSeq™-C0050 anti-human CD19 Antibody | HIB19 | BioLegend | 302265 |
| TotalSeq™-C0047 anti-human CD56 (NCAM) Antibody | 5.1H11 | BioLegend | 362559 |
| TotalSeq™-C0046 anti-human CD8 Antibody | SK1 | BioLegend | 344753 |
| TotalSeq™-C0053 anti-human CD11c Antibody | S-HCL-3 | BioLegend | 371521 |
| TotalSeq™-C0081 anti-human CD14 Antibody | M5E2 | BioLegend | 301859 |
| TotalSeq™-C0083 anti-human CD16 Antibody | 3G8 | BioLegend | 302065 |
| TotalSeq™-C0087 anti-human CD45RO Antibody | UCHL1 | BioLegend | 304259 |
| TotalSeq™-C0088 anti-human CD279 (PD-1) Antibody | EH12.2H7 | BioLegend | 329963 |
| TotalSeq™-C0169 anti-human CD366 (Tim-3) Antibody | F38-2E2 | BioLegend | 345049 |
| TotalSeq™-C0151 anti-human CD152 (CTLA-4) Antibody | BNI3 | BioLegend | 369621 |
| TotalSeq™-C0804 anti-human CD186 (CXCR6) Antibody | K041E5 | BioLegend | 356023 |
| TotalSeq™-C0159 anti-human HLA-DR Antibody | L243 | BioLegend | 307663 |
| TotalSeq™-C0423 anti-human MERTK Antibody | 590H11G1E3 | BioLegend | 367623 |
| TotalSeq™-C0148 anti-human CD197 (CCR7) Antibody | G043H7 | BioLegend | 353251 |
| TotalSeq™-C0141 anti-human CD195 (CCR5) Antibody | J418F1 | BioLegend | 359137 |
| TotalSeq™-C0358 anti-human CD163 Antibody | GHI/61 | BioLegend | 333637 |
| TotalSeq™-C0576 anti-human CD49d Antibody | 9F10 | BioLegend | 304345 |
| TotalSeq™-C0091 Mouse IgG2a, κ isotype Ctrl Antibody | MOPC-173 | BioLegend | 400293 |
| TotalSeq™-C0092 Mouse IgG2b, κ isotype Ctrl Antibody | MCP-11 | BioLegend | 400381 |
| TotalSeq™-C0090 Mouse IgG1, κ isotype Ctrl Antibody | MOPC-21 | BioLegend | 400187 |

***Table S13 Inclusion and exclusion criteria of PREDICT study*** (1)

| Inclusion criterion | 1. The patient is admitted/referred to study center with AD of cirrhosis (ascites, overt encephalopathy, new onset of non-obstructive jaundice, GI-hemorrhage and/or bacterial infections), but without ACLF (as defined according to the CANONIC study) at study inclusion. ACLF patients at study inclusion will serve as positive controls. |
| --- | --- |
| Exclusion criteria | 1. Pregnancy. 2. Age <18 years. 3. Patients with acute or subacute liver failure without underlying cirrhosis. 4. Patients with cirrhosis who develop decompensation in the postoperative period following partial hepatectomy. 5. Evidence of current malignancy except for non-melanocytic skin cancer and hepatocellular carcinoma within Milan criteria. 6. Presence or history of severe extra-hepatic diseases (e.g., chronic renal failure requiring hemodialysis, severe heart disease (NYHA > II), severe chronic pulmonary disease (GOLD > III), severe neurological and psychiatric disorders) 7. HIV-positive patients. 8. Previous liver or other transplantation. 9. Admission/referral of more than 72 hours before inclusion. 10. Patients who decline to participate or cannot provide prior written informed consent and when there is documented evidence that the patient has no legal surrogate decision maker, and it appears unlikely that the patient will regain consciousness or sufficient ability to provide delayed informed consent. 11. Physician’s denial (e.g. the investigator considers that the patient will   not follow the protocol scheduled). |

***Table S14 Inclusion and exclusion criteria of ACLARA study*** **(2)**.

| Inclusion criterion | 1. Consecutive patients with cirrhosis admitted to hospital for more than one day for the treatment of acute decompensation of cirrhosis as defined by one or more of the following complications: ascites, hepatic encephalopathy, GI-bleeding and/or bacterial or fungal infections (only in cases with prior decompensated cirrhosis). |
| --- | --- |
| Exclusion criteria | 1. Pregnancy. 2. Age <18 years. 3. More than 4 days between admission to the Unit and inclusion into the study. 4. Patients with acute or subacute liver failure without underlying cirrhosis. 5. Patients with cirrhosis admitted for more than one day for scheduled procedures (e.g., band ligation, transjugular intrahepatic portosystemic shunting). 6. Outpatients with cirrhosis and refractory ascites who stay at hospital >1 day for scheduled treatments (i.e., large-volume paracentesis, or variceal banding). 7. Patients with cirrhosis who develop decompensation in the postoperative period following partial hepatectomy 8. Evidence of current locally advanced or metastatic malignancy (patients with hepatocellular carcinoma within the Milan criteria, non-melanocytic skin cancer and controlled breast or prostate cancer can be included) 9. Previously known severe extra-hepatic diseases (e.g., chronic renal failure requiring hemodialysis, severe heart disease; severe chronic pulmonary disease, psychiatric disorders). 10. Immunosuppressive drugs other than corticosteroids at a dose for severe alcoholic hepatitis, unless the patient has autoimmune cirrhosis. 11. HIV infection. 12. Patients who cannot provide prior informed consent and when there is documented evidence that the patient has no legal surrogate decision maker, and it appears unlikely that the patient will regain consciousness or sufficient ability to provide delayed informed consent, physician and team not committed to intensive care if needed. 13. Previous liver transplantation |

**Supplementary Figures**

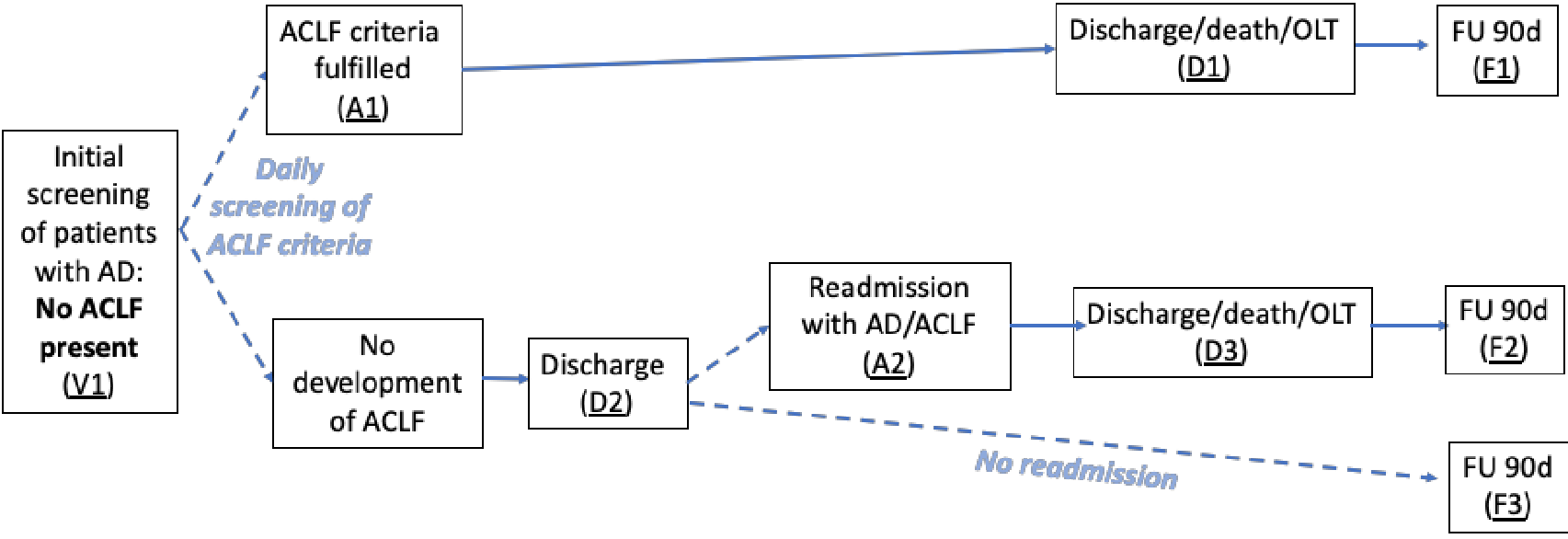

***Figure S1. Study design of the single-cell study cohort.***

*Abbreviations*: V, visit; A, admission; D, discharge; F, follow-up visit; OLT, orthotopic liver transplant; AD: acute decompensation; ACLF: acute-on-chronic liver failure; OLT, orthotopic liver transplant; FU, follow-up.

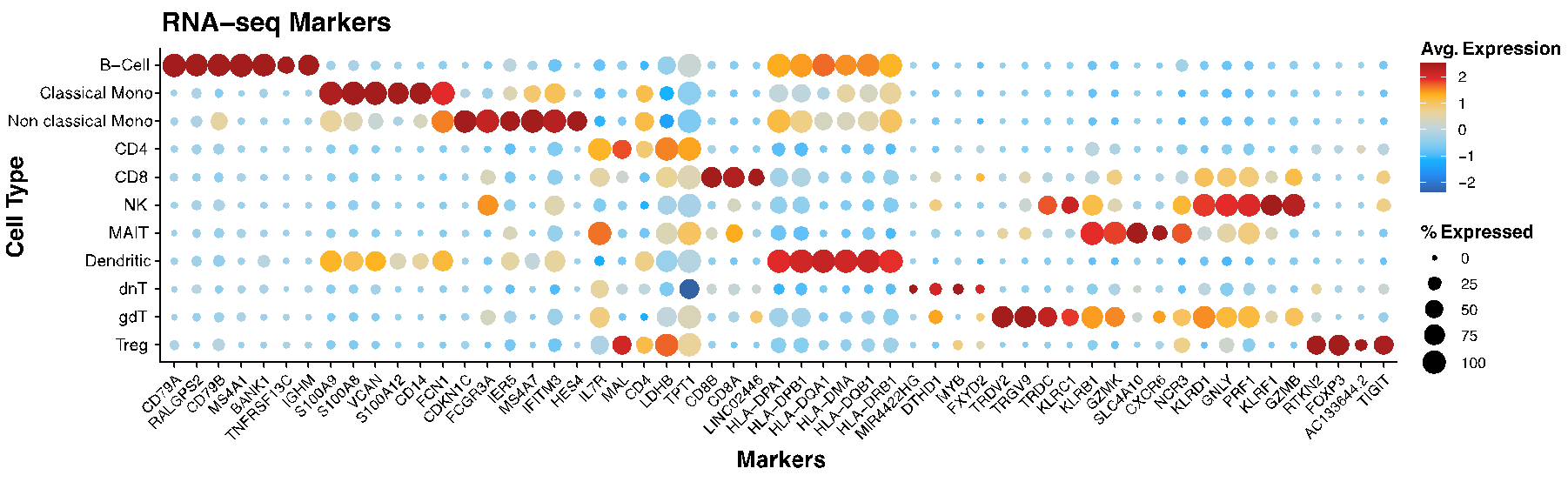

***Figure S2. Gene expression markers used for annotating the single-cell cohort clusters.***

Dot plot displaying the average expression levels and percentage of cells with the top gene expression markers across the different PBMC cell types. The color scale indicates the average scaled expression level of each marker (blue = low, red = high). The dot size reflects the percentage of cells within each cluster that express the marker. *Abbreviations*: Avg, average.

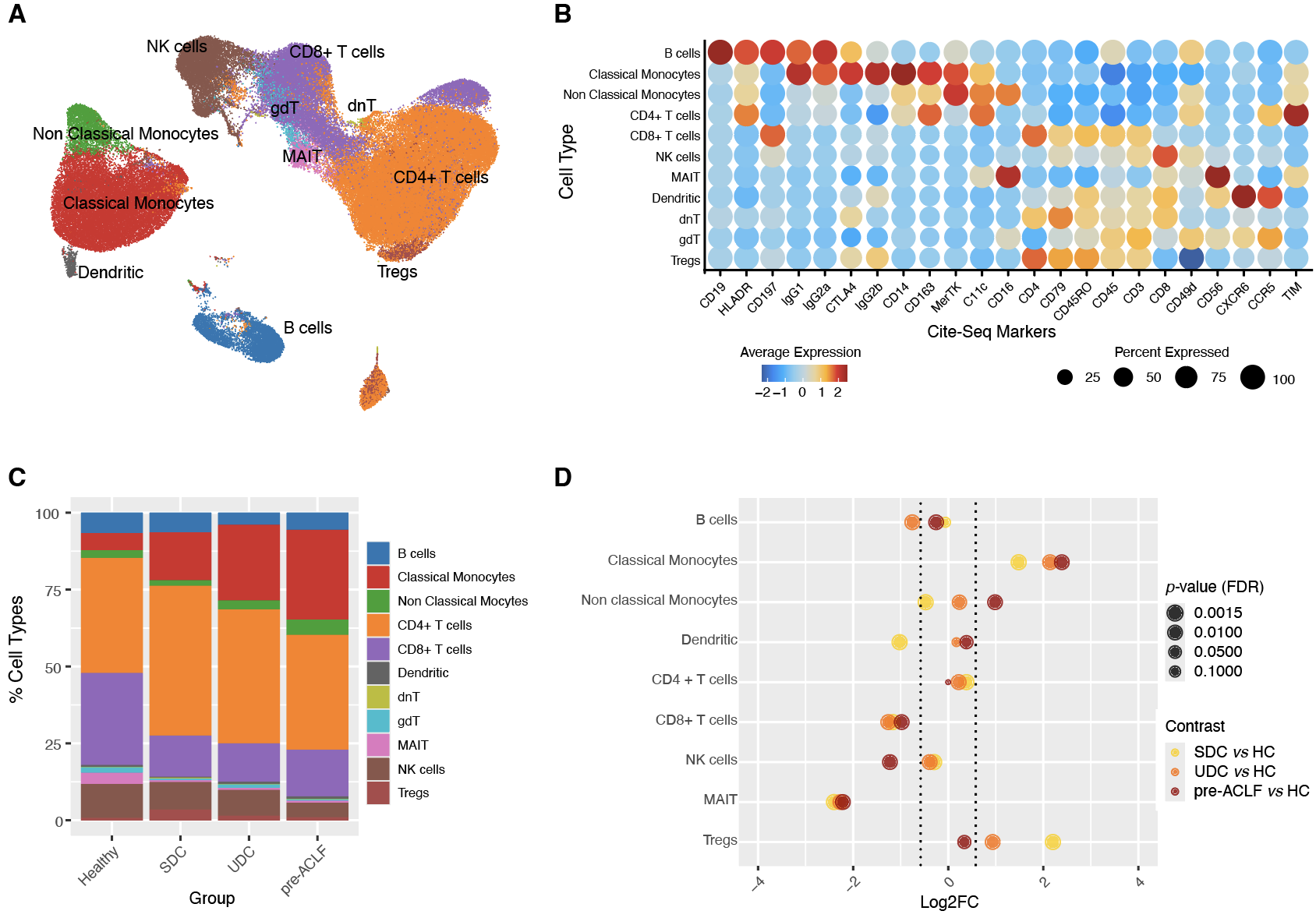

***Figure S3. Single-cell epitope profiling of the single-cell study cohort.***

**(A)** UMAP projection of epitope profiling data derived from CITE-seq of the PBMCs from all groups, colored by cell type. **(B)** The surface marker expression dot plot shows the average scaled expression and percentage of cells expressing specific surface markers for each identified cell type. The color scale indicates the average expression level of each marker (blue = low, red = high). The dot size reflects the percentage of cells within each cluster that express the marker. **(C)** Stacked bar plots showing the proportions of each immune cell type within PBMC samples from HC, SDC, UDC, and pre-ACLF groups. **(D)** Differences in cell type proportions**:** Cleveland dot plot displaying the log2 fold-change in cell type proportions between SDC vs. HC, UDC vs. HC, and pre-ACLF vs. HC. The size of the dots indicates the significance (adjusted p-value) of the differences observed. The plot highlights significant increases in classical monocytes and non-classical monocytes in pre-ACLF patients compared to other groups. Log2FC and FDR were estimated, respectively, using bootstrapping and permutation analysis. *Abbreviations*: Avg, average; AD, acute decompensation; classical Mono, classical monocytes; non-classical Mono, non-classical monocytes; CD4, CD4+ T cells; CD8, CD8+ T cells; dnT, double negative T cells; FDR, false discovery rate; gdT, gamma delta T cells; HC, healthy controls; NK cells, natural killer cells; MAIT cells, mucosal-associated invariant T cells; SDC, stable decompensated cirrhosis; pre-ACLF, pre-acute-on-chronic liver failure; Tregs, regulatory T cells; UDC, unstable decompensated cirrhosis.

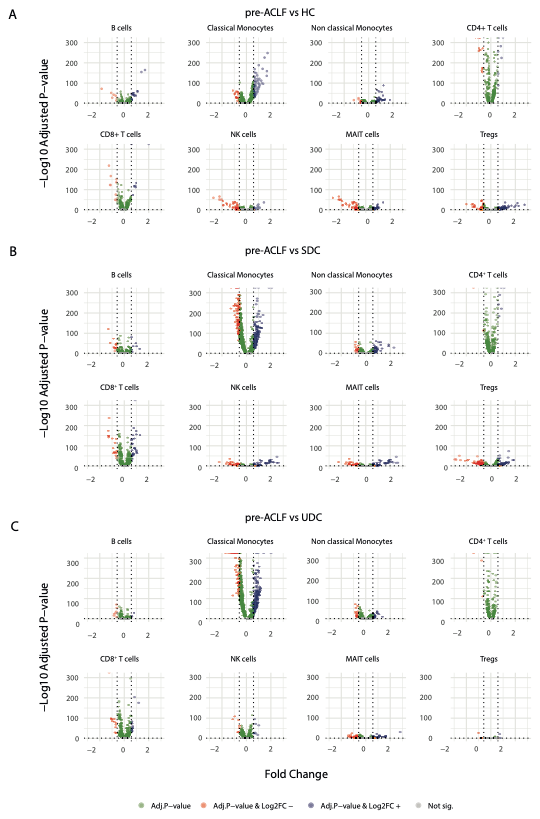

***Figure S4. Volcano plots representing the fold change (log2 FC) and statistical significance (adjusted p-value) of differentially expressed genes (DEGs) for different immune cell populations comparing healthy controls and patients with SDC, UDC, and pre-ACLF.*** Each plot represents the -Log10 of the adjusted p-value against the fold-change (Log2FC) of gene expression, with different colors indicating the significance and direction of the fold-change. Green dots: Genes with significant adjusted p-value only. Red dots: Genes with significant adjusted p-value and negative fold-change (downregulated in UDC). Blue dots: Genes with significant adjusted p-value and positive fold-change (upregulated in UDC). Grey dots: Genes not showing significant differential expression. **(A)** illustrates the comparison of pre-ACLF against HC, **(B)** illustrates the comparison of pre-ACLF against SDC and **(C)** illustrates the comparison of pre-ACLF against UDC. *Abbreviations:* AD, acute decompensation; HC, healthy controls; NK cells, natural killer cells; MAIT cells, mucosal-associated invariant T cells; PBMCs, peripheral blood mononuclear cells; pre-ACLF, pre-acute-on-chronic liver failure; SDC, stable decompensated cirrhosis; Tregs, regulatory T cells; UDC, unstable decompensated cirrhosis.

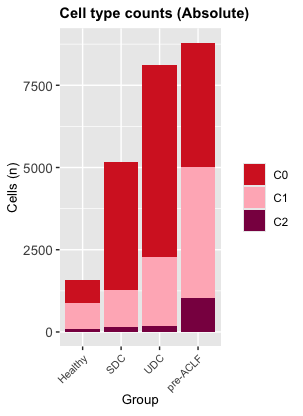

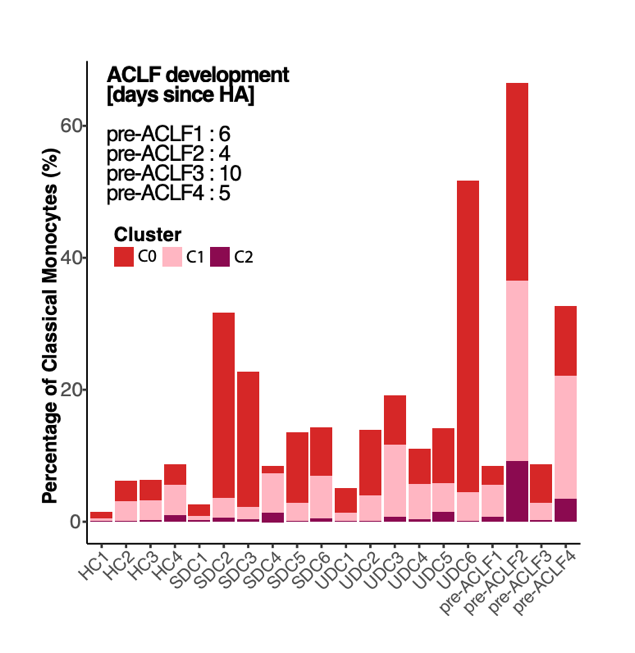

**B**

**A**

***Figure S5.*** ***Distribution of monocyte subclusters across the study groups and patients.*** *(A) Absolute counts of each monocyte subcluster in each study group. (B) Relative percentages of each monocyte subcluster in each individual patient.*

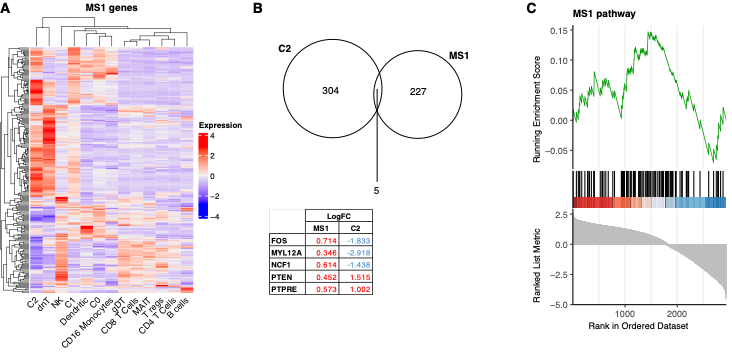

***Figure S6. Comparison of C2 against MS1 monocytes.* (A)** The heatmap shows the expression data of the MS1 genes across the different cell types in our single cell data. The color scale (from purple to red) represents the mean expression value, where dark red indicates high expression and dark purple indicates low expression. **(B)** The Venn diagram (top) displays the intersection of the differentially expressed genes in the C2 cluster and the genes reported in the MS1 monocytes. There is an overlap of 5 genes, whose expression differences are shown in the table (bottom). **(C)** Gene Set Enrichment Plot showing the ranking of MS1 genes based on C2 expression. The plot reveals no significant enrichment, as the genes are uniformly distributed across the dataset.

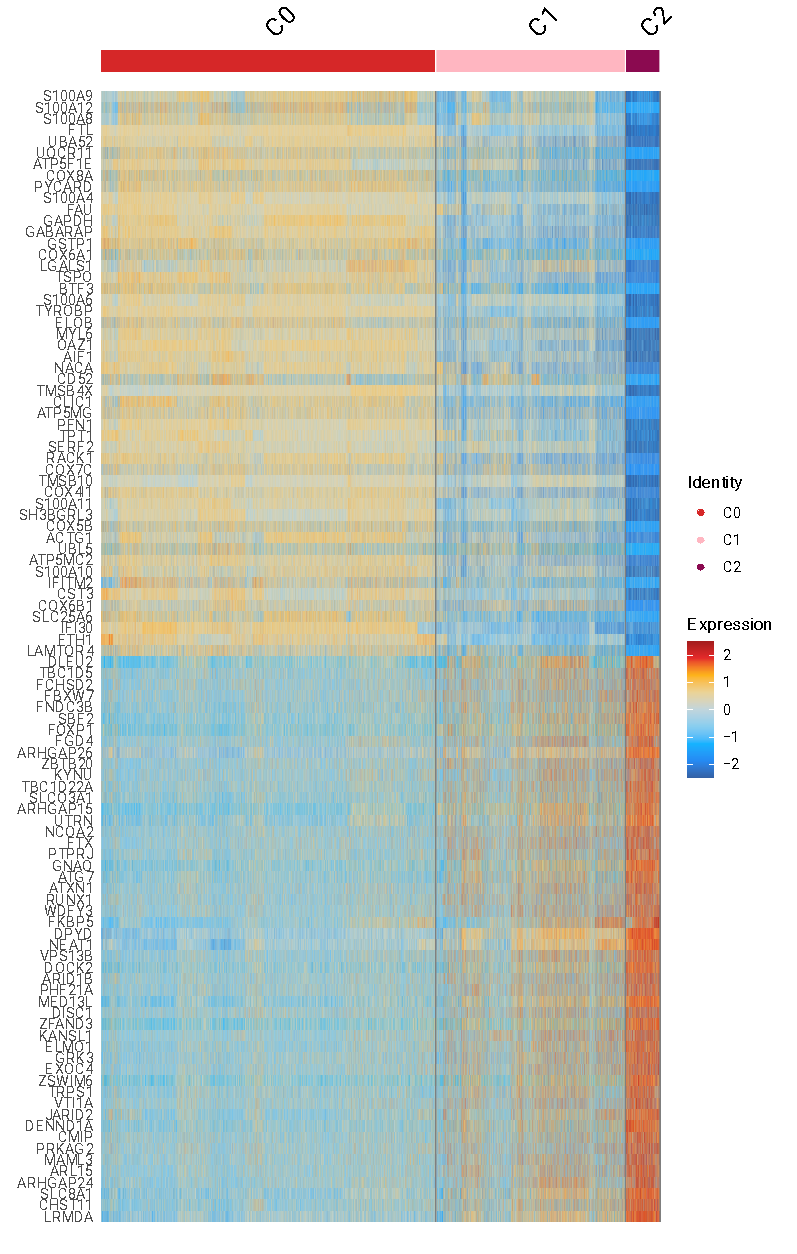

***Figure S7. Heatmap showing the top 50 up and 50 down-regulated genes comparing C0, C1, and C2 monocytes.*** Heatmap displaying the scaled expression values of the top 50 up and 50 down-regulated in C0, C1, and C2 monocytes.

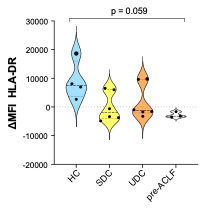

***Figure S8. FACS analysis of HLA-DR expression on classical monocytes of single-cell study cohort.*** Violin plots illustrating the distribution, median, and IQR of the ΔMIF (subtraction of the mean MFI count in the SDC group from single MFI counts in the four study groups) of surface HLA-DR expression on classical monocytes. Samples were drawn from the single-cell study cohort, including healthy controls (n=4) and patients with SDC (n=6), UDC (n=6), and pre-ACLF (n=4). The Wilcoxon-Mann-Whitney test was used to test statistically significant differences in relative HLA-DR expression within the four study groups. *Abbreviations:* HC, healthy controls; MIF, delta of mean fluorescence intensity; pre-ACLF, pre-acute-on-chronic liver failure; SDC, stable decompensated cirrhosis; UDC, unstable decompensated cirrhosis.

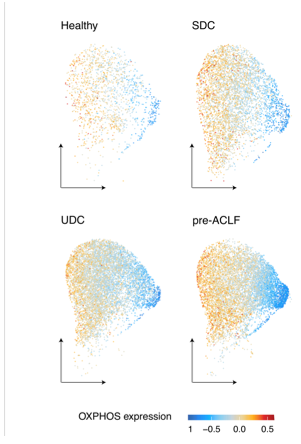

***Figure S9. OXPHOS expression distribution.*** UMAP restricted to classical monocytes showing OXPHOS signature scores by patient group (Healthy, SDC, UDC, pre-ACLF). *Abbreviations:* OXPHOS, oxidative phosphorylation; pre-ACLF, pre-acute-on-chronic liver failure; SDC, stable decompensated cirrhosis; UDC, unstable decompensated cirrhosis.

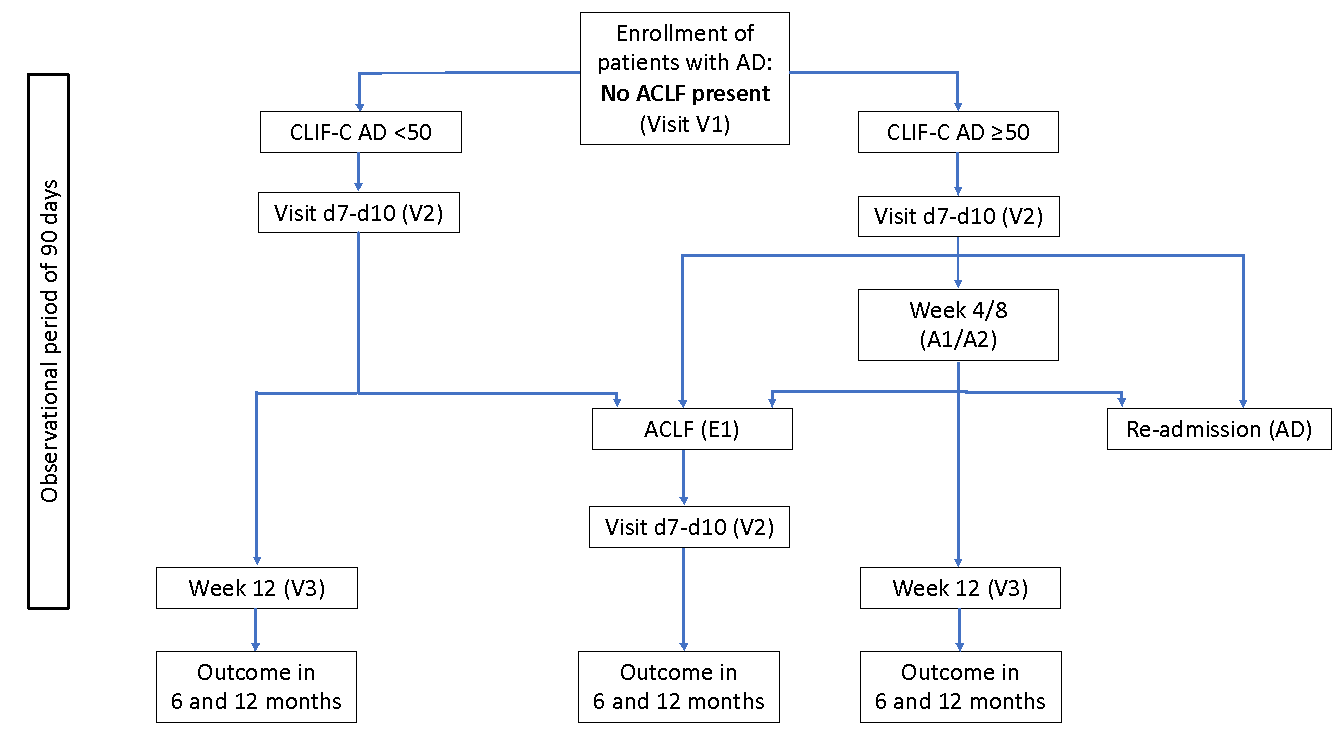

***Figure S10. Study design PREDICT study (1).*** *Explanations*: V, planned visit; E, visit in case of ACLF; A, additional planned visit; AD, readmission visit.

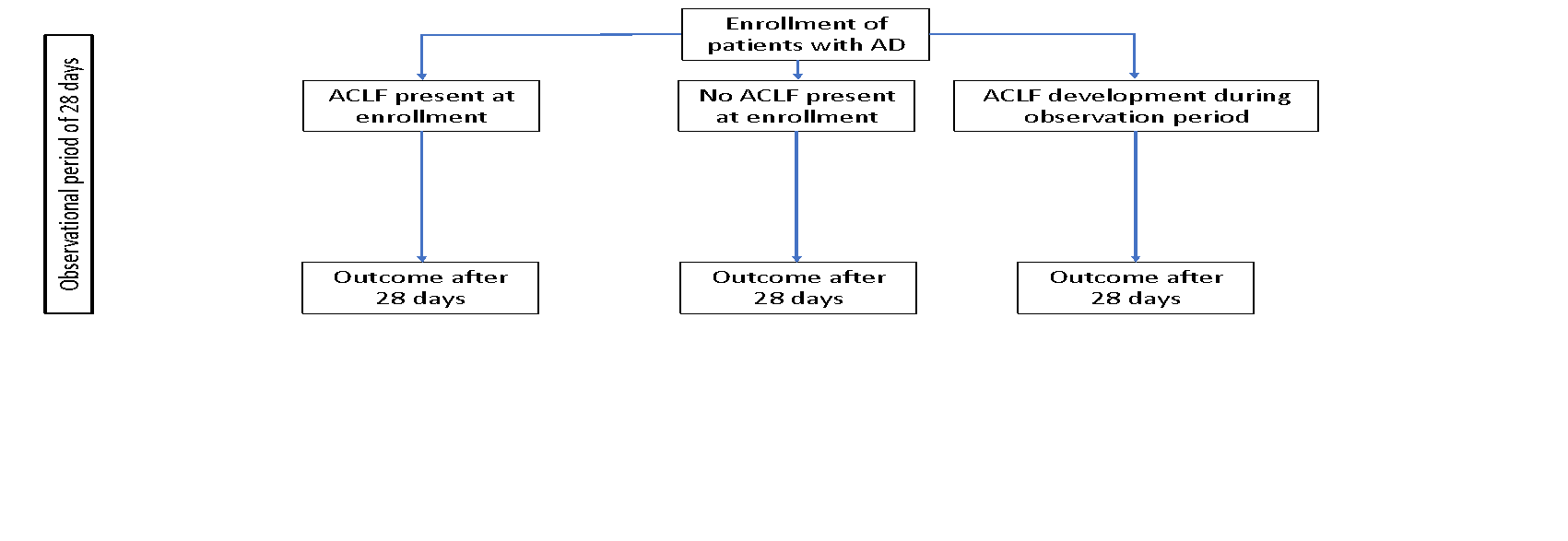

***Figure S11. Study design ACLARA study (2).***

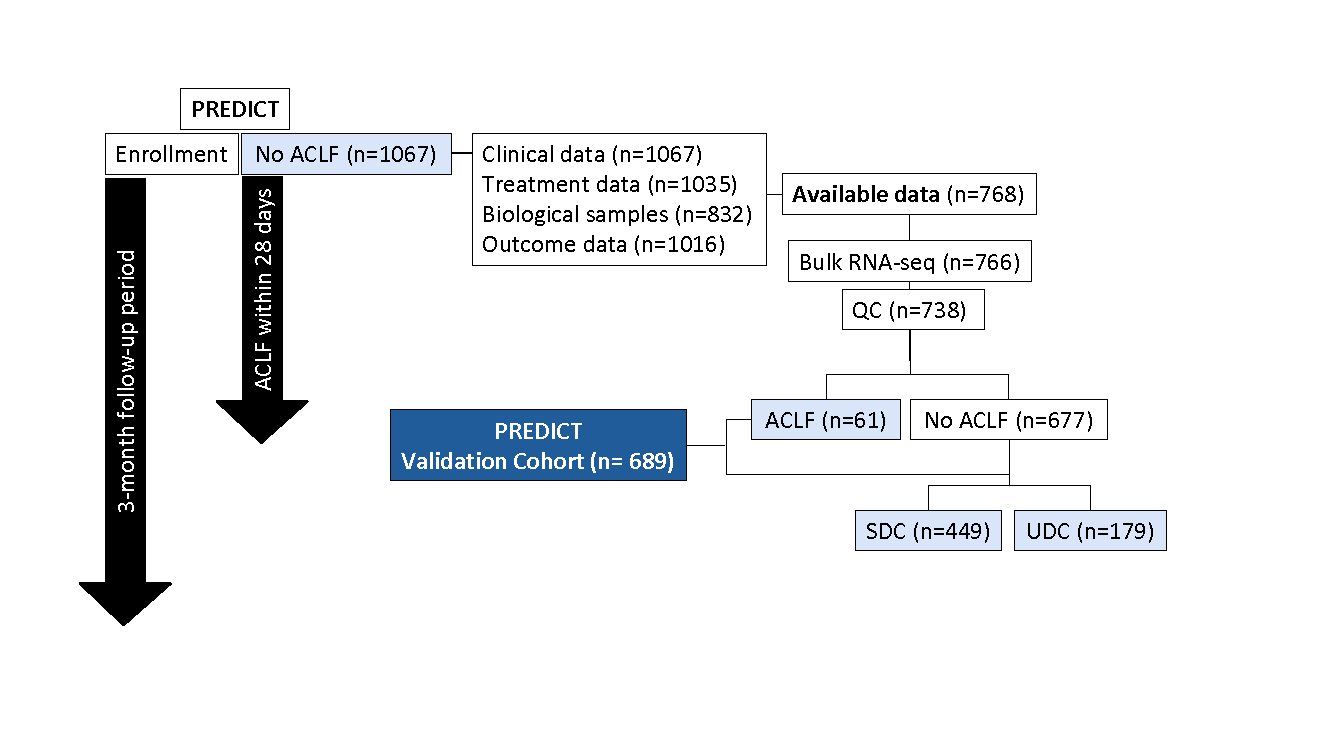

***Figure S12. Flow chart for patient selection from PREDICT cohort (1).***

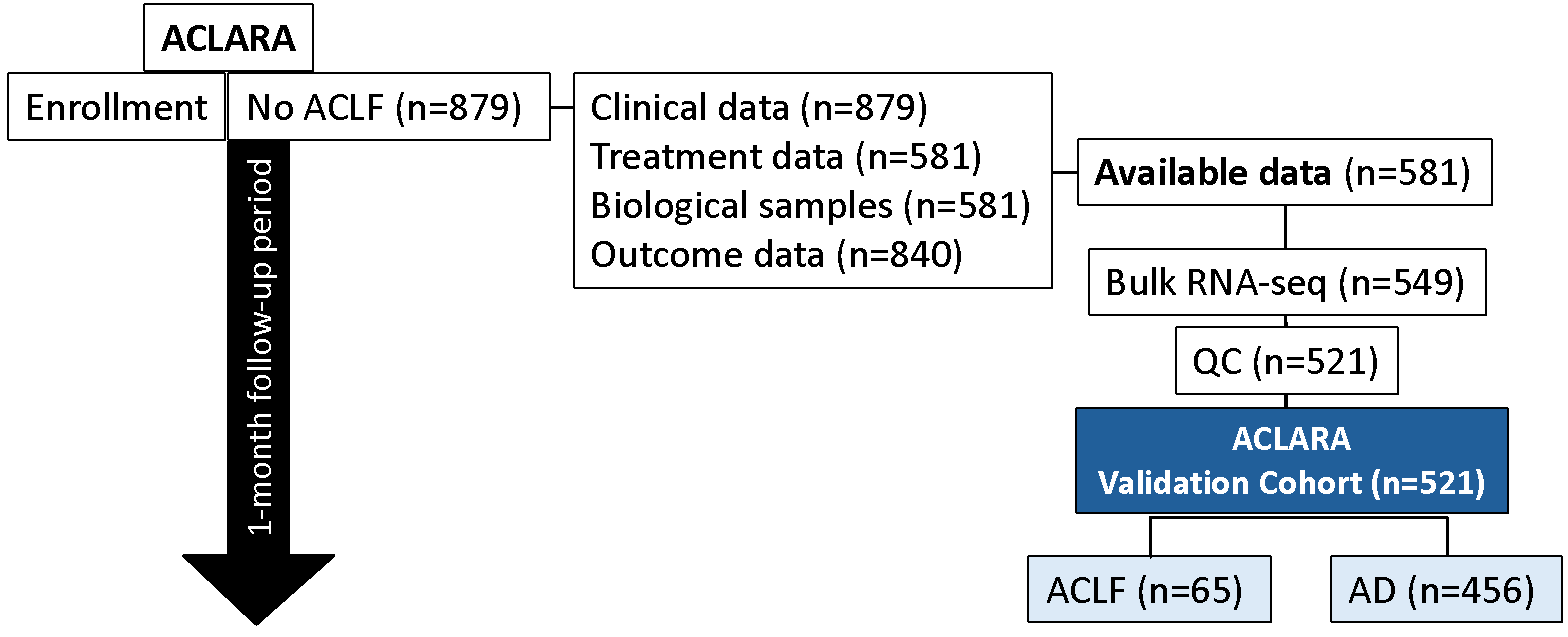

***Figure S13. Flow chart for patient selection from ACLARA cohort (2).***

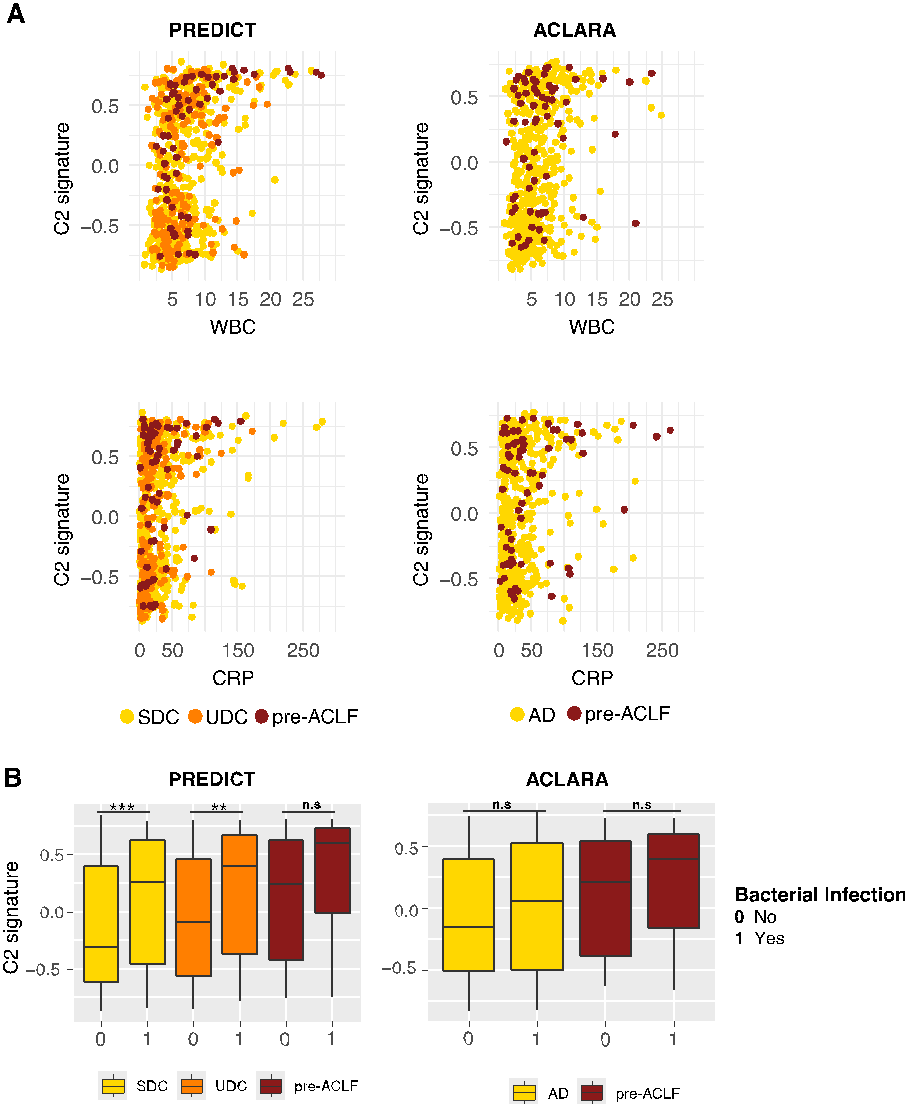

***Figure S14. C2 signature average expression by group and bacterial infection.***

**(A)** Correlation of C2 signature values with white blood cell (WBC) count and C-reactive protein (CRP) levels across different patient groups in PREDICT (left) and ACLARA (right) cohorts. Top row: C2 signature vs. WBC count. Bottom row: C2 signature vs. CRP levels. Each point represents an individual, colored by patient group: SDC, yellow; UDC, orange; pre-ACLF, dark red for the PREDICT cohort, and AD, yellow and pre-ACLF, dark red for the ACLARA cohort. **(B)** Boxplots showing the C2 module scores for patients with no bacterial infection (0) and bacterial infection (1) across different groups in the PREDICT cohort (1) (left) and in the ACLARA cohort (2) (right). Wilcoxon Signed-Rank Test was used to assess the differences of the mean between groups ** adjusted p-value < 0.01; * adjusted p-value < 0.01. *Abbreviations:* AD, acute decompensation; SDC, stable decompensated cirrhosis; UDC, unstable decompensated cirrhosis; pre-ACLF, pre-acute-on-chronic liver failure; WBC, white blood cell count; CRP, C-reactive protein.

20.	R Core Team. R: A language and environment for statistical computing. Vienna, Austria: R Foundation for Statistical Computing; 2021.
